# Repeat Subcutaneous Administration of REGEN-COV^®^ in Adults is Well-Tolerated and Prevents the Occurrence of COVID-19

**DOI:** 10.1101/2021.11.10.21265889

**Authors:** Flonza Isa, Eduardo Forleo-Neto, Jonathan Meyer, Wenjun Zheng, Scott Rasmussen, Danielle Armas, Masaru Oshita, Cynthia Brinson, Steven Folkerth, Lori Faria, Ingeborg Heirman, Neena Sarkar, Bret J. Musser, Shikha Bansal, Meagan P. O’Brien, Kenneth C. Turner, Samit Ganguly, Adnan Mahmood, Ajla Dupljak, Andrea T. Hooper, Jennifer D. Hamilton, Yunji Kim, Bari Kowal, Yuhwen Soo, Gregory P. Geba, Leah Lipsich, Ned Braunstein, George D. Yancopoulos, David M. Weinreich, Gary A. Herman, the COVID-19 Multi-dose Trial Team

## Abstract

**Background:** Data show that a single dose of casirivimab and imdevimab (REGEN-COV^®^) is effective in treating hospitalized individuals and outpatients with COVID-19 and in post-exposure prophylaxis. We present results from a phase 1, double-blind, placebo-controlled trial evaluating the safety, tolerability, and efficacy of repeat monthly doses of subcutaneous (SC) REGEN-COV in uninfected adult volunteers who were healthy or had chronic stable medical conditions.

**Methods:** Subjects were randomized (3:1) to SC REGEN-COV 1200 mg or placebo dosed every 4 weeks for up to 6 doses. The primary and secondary endpoints evaluated the safety, pharmacokinetics, and immunogenicity of multiple-dose administration of REGEN-COV. Efficacy was evaluated by the incidence of COVID-19 or SARS-CoV-2 seroconversion.

**Results:** In total, 969 subjects were treated. Repeat monthly dosing of SC REGEN-COV led to a 92.4% relative risk reduction in clinically-defined COVID-19 compared to placebo (3/729 [0.4%] vs 13/240 [5.4%]; odds ratio: 0.07 [95% CI, 0.01–0.27]), and a 100% reduction in laboratory-confirmed COVID-19 (0/729 vs 10/240 [4.2%]; odds ratio 0.00). Development of anti-drug antibodies was low (<5% subjects). No grade ≥3 injection-site reactions (ISRs) or hypersensitivity reactions were reported. A slightly higher percentage of subjects reported TEAEs with REGEN-COV (54.9%) than placebo (48.3%), due to ISRs (all grade 1-2). Serious adverse events were rare and occurred at similar percentages in the REGEN-COV and placebo groups. No deaths were reported in the 6-month treatment period.

**Conclusions:** Repeated monthly administration of 1200 mg SC REGEN-COV was well-tolerated with low immunogenicity, and showed a substantial risk reduction in COVID-19 occurrence.

(ClinicalTrials.gov identifier, NCT04519437)

## Introduction

Severe acute respiratory syndrome coronavirus 2 (SARS-CoV-2) is the causal agent of coronavirus disease 2019 (COVID-19),^1-3^ which emerged in December 2019 and was declared a global pandemic in March 2020.^4-6^ Other than vaccines, there are currently no approved therapies that can be used for chronic prevention of COVID-19.^7,8^ Such treatments are urgently needed, especially in populations who are immunocompromised or have a medical condition or other reason making them unlikely to respond to or be protected by vaccination.^9^

Casirivimab and imdevimab (REGEN-COV^®^) is a combination of distinct neutralizing monoclonal antibodies that simultaneously bind non-overlapping epitopes of the receptor binding domain of the SARS-CoV-2 spike protein, thereby preventing the virus from infecting host cells.^10-12^ Preclinical and human studies indicate that administering these 2 antibodies in combination provides potent neutralization against currently circulating viral variants of concern including beta, gamma, epsilon, and delta variants, and may protect against the selection of new treatment-resistant SARS-CoV-2 variants.^10,13^ Previous trials with REGEN-COV have shown clinical benefit in the treatment setting, reducing the likelihood of hospitalization and death by over 70% in high-risk SARS-CoV-2 infected outpatients,^14,15^ and by reducing mortality in seronegative hospitalized patients by 21%.^16^ Additionally, a single dose of REGEN-COV reduced both infection and symptomatic COVID-19 in close contacts of infected individuals.^17^ On the basis of these studies, REGEN-COV is currently authorized for treatment and post-exposure prophylaxis of COVID-19 in certain settings in the US,^18,19^ and for treatment and prevention of COVID-19 in other jurisdictions (known as Ronapreve™ outside of the US).^20^ This study evaluated the safety, tolerability, and efficacy of monthly dosing of subcutaneous (SC) REGEN-COV in uninfected individuals.

## Methods

### Study Design and Treatment

This phase 1, double-blind, placebo-controlled, study was conducted at 7 sites in the US (clinicaltrials.gov identifier, NCT04519437). The study was comprised of a screening/baseline period (up to 7 days), a treatment period (up to 24 weeks), and a follow-up period (28 weeks) (**Supplementary Figure 1**). Subjects underwent an end-of-treatment period visit 1 month after their final dose of study drug, before entering the 28-week follow-up period leading to an end of study visit. As of the data cut-off of May 21, 2021, all eligible subjects had completed the end-of-treatment visit after receipt of up to 6 doses of study drug; follow-up was ongoing, with not all subjects having completed the entire study. The rationale for 1200 mg SC dose selection and methods for dose administration are provided in the **Supplementary Appendix**.

### Subjects

Eligible subjects were uninfected adult volunteers, aged 18–90 years, who were healthy or had chronic but stable medical conditions, without signs/symptoms suggestive of COVID-19, and who were confirmed to be SARS-CoV-2 negative by central lab reverse transcription polymerase chain reaction (RT-PCR) of nasopharyngeal swab ≤72 hours of randomization. While in the treatment period, subjects who met any of the following criteria were discontinued from study drug and were moved into the follow-up period: tested positive for SARS-CoV-2, developed symptomatic COVID-19, experienced an adverse event (AE) that led to study drug discontinuation, received a dose of an investigational COVID-19 vaccine, or were unblinded per protocol to receive a COVID-19 vaccine. A full list of inclusion and exclusion criteria as well as diagnostic criteria for COVID-19 are presented in the **Supplementary Appendix**.

### COVID-19 Vaccination During the Study

Subjects who elected to receive COVID-19 vaccination were discontinued from study drug and entered follow-up. Handling of COVID-19 vaccination during the study is described in the **Supplementary Appendix**.

### Outcome Measures

The primary endpoints were to assess the incidence of AEs of special interest (AESIs), defined as grade ≥3 injection-site reactions (ISRs) or hypersensitivity reactions, that occurred within 4 days of administration of REGEN-COV or placebo, as well as the concentrations of REGEN-COV in serum over time. All AEs were graded for severity using the National Cancer Institute-Common Terminology Criteria for Adverse Events (NCI-CTCAE) v. 5.0. Investigators assessed AEs to determine seriousness, relatedness to investigational product, and whether AESI criteria were met.

Secondary endpoints included: the proportion of subjects with treatment-emergent AEs (TEAEs), the proportion of subjects who achieved or exceeded the target concentration of REGEN-COV in serum (20 µg/mL) at the end of each 4-week dosing interval, and immunogenicity (measured by anti-drug antibodies [ADAs]) to REGEN-COV.

Exploratory efficacy endpoints assessed the incidence and severity of symptomatic SARS-CoV-2 infection (ie, COVID-19) during the treatment and follow-up periods and the proportion of baseline anti-SARS-CoV-2 seronegative subjects who converted to seropositive for SARS-COV-2 anti-nucleocapsid IgG antibodies post-baseline; seroconversion from negative to positive for SARS-COV-2 anti-nucleocapsid IgG antibodies was considered indicative of an incident SARS-CoV-2 infection. Diagnosis of COVID-19 and assessment of SARS-CoV-2 serostatus are described in the **Supplementary Appendix**.

### Statistical Analysis

The study aimed to enroll approximately 940 subjects (705 and 235 subjects in the REGEN-COV and placebo groups, respectively) to ensure sufficient safety data to support multiple dose administration of REGEN-COV.

Treatment compliance/administration, the exploratory efficacy endpoints, and all clinical safety variables were analyzed in the safety analysis set, which was comprised of all randomized subjects who received any study drug. Subjects who received ≥1 dose of REGEN-COV were classified in the REGEN-COV group. The pharmacokinetic (PK) analysis set included all subjects who received any study drug and had ≥1 non-missing post-baseline result.

Summary statistics were provided for safety analyses, exploratory efficacy analyses (with nominal *P*-values) and PK and immunogenicity variables. Handling of missing data and PK analysis methods are described in the **Supplementary Appendix**.

### Study Oversight

Study oversight is described in the **Supplementary Appendix**.

## Results

### Demographics and Baseline Characteristics

Baseline characteristics were balanced between the REGEN-COV and placebo groups. The overall median age was 48 years, 44.9% of subjects were female, 10% of subjects identified as African American, and 23.4% identified as Hispanic or Latino (**Table 1**). There were 6 (0.6%) subjects who tested positive for SARS-CoV-2 via a centralized RT-PCR assay at baseline who had previously tested negative at screening with a local test; these subjects received 1 dose of study drug prior to the availability of the RT-PCR result from the central lab and were discontinued from additional doses of study drug and excluded from efficacy analyses but continued to be followed for safety. Subjects were included in the study regardless of SARS-CoV-2 antibody serostatus. At baseline, 825 (85.1%) subjects were seronegative for both SARS-CoV-2 anti-spike and anti-nucleocapsid antibodies, 101 (10.4%) were seropositive for anti-spike and/or anti-nucleocapsid antibodies, and 43 (4.4%) were of borderline or unknown serostatus (**Table 1**). Medical history was comparable between the REGEN-COV and placebo groups with 20% of subjects reporting ≥1 medical condition considered high-risk for progression to severe COVID-19;^21-23^ the most common of these were hypertension and asthma (**Table 1; Supplementary Table 1**).

**Table 1.**
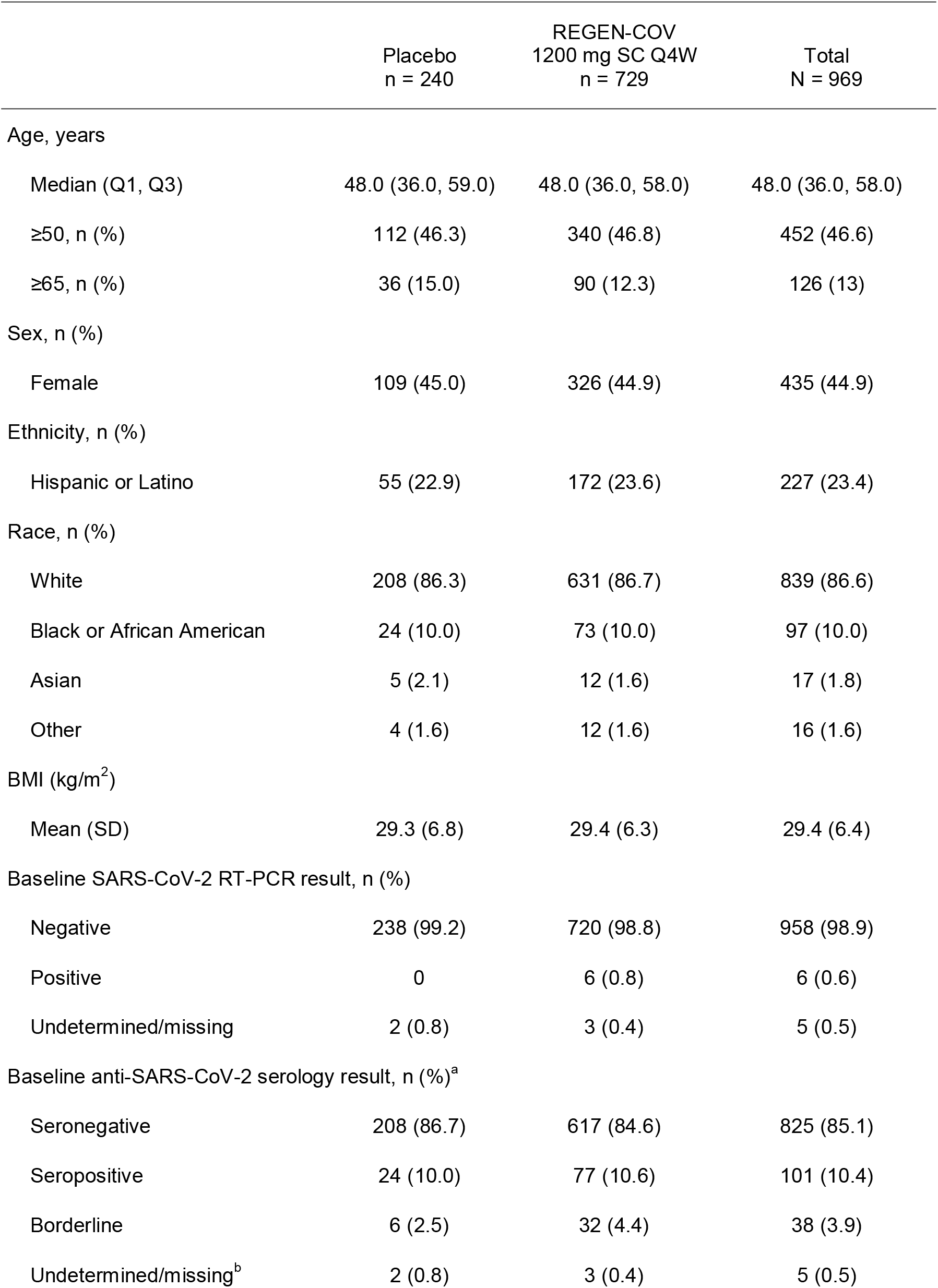
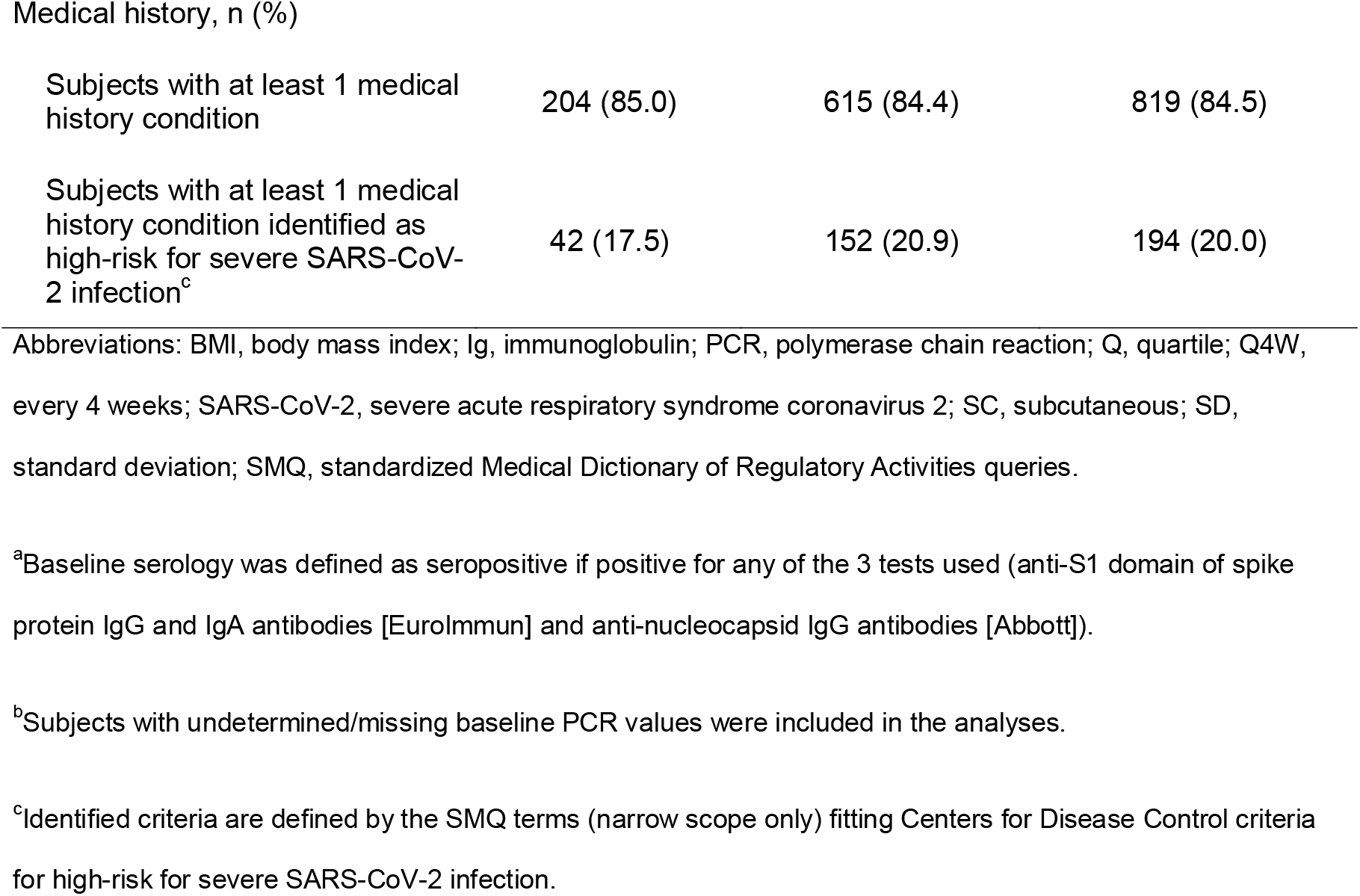
Demographics and Baseline Characteristics.

|  | Placebo<br>n = 240 | REGEN-COV<br>1200 mg SC Q4W<br>n = 729 | Total<br>N = 969 |
| --- | --- | --- | --- |
| Age, years |  |  |  |
| Median (Q1, Q3) | 48.0 (36.0, 59.0) | 48.0 (36.0, 58.0) | 48.0 (36.0, 58.0) |
| ≥50, n (%) | 112 (46.3) | 340 (46.8) | 452 (46.6) |
| ≥65, n (%) | 36 (15.0) | 90 (12.3) | 126 (13) |
| Sex, n (%) |  |  |  |
| Female | 109 (45.0) | 326 (44.9) | 435 (44.9) |
| Ethnicity, n (%) |  |  |  |
| Hispanic or Latino | 55 (22.9) | 172 (23.6) | 227 (23.4) |
| Race, n (%) |  |  |  |
| White | 208 (86.3) | 631 (86.7) | 839 (86.6) |
| Black or African American | 24 (10.0) | 73 (10.0) | 97 (10.0) |
| Asian | 5 (2.1) | 12 (1.6) | 17 (1.8) |
| Other | 4 (1.6) | 12 (1.6) | 16 (1.6) |
| BMI (kg/m<sup>2</sup>) |  |  |  |
| Mean (SD) | 29.3 (6.8) | 29.4 (6.3) | 29.4 (6.4) |
| Baseline SARS-CoV-2 RT-PCR result, n (%) |  |  |  |
| Negative | 238 (99.2) | 720 (98.8) | 958 (98.9) |
| Positive | 0 | 6 (0.8) | 6 (0.6) |
| Undetermined/missing | 2 (0.8) | 3 (0.4) | 5 (0.5) |
| Baseline anti-SARS-CoV-2 serology result, n (%)<sup>a</sup> |  |  |  |
| Seronegative | 208 (86.7) | 617 (84.6) | 825 (85.1) |
| Seropositive | 24 (10.0) | 77 (10.6) | 101 (10.4) |
| Borderline | 6 (2.5) | 32 (4.4) | 38 (3.9) |
| Undetermined/missing<sup>b</sup> | 2 (0.8) | 3 (0.4) | 5 (0.5) |
| Medical history, n (%) |  |  |  |
| Subjects with at least 1 medical history condition | 204 (85.0) | 615 (84.4) | 819 (84.5) |
| Subjects with at least 1 medical history condition identified as high-risk for severe SARS-CoV-2 infection<sup>c</sup> | 42 (17.5) | 152 (20.9) | 194 (20.0) |

Abbreviations: BMI, body mass index; Ig, immunoglobulin; PCR, polymerase chain reaction; Q, quartile; Q4W, every 4 weeks; SARS-CoV-2, severe acute respiratory syndrome coronavirus 2; SC, subcutaneous; SD, standard deviation; SMQ, standardized Medical Dictionary of Regulatory Activities queries.
<sup>a</sup>Baseline serology was defined as seropositive if positive for any of the 3 tests used (anti-S1 domain of spike protein IgG and IgA antibodies [EuroImmun] and anti-nucleocapsid IgG antibodies [Abbott]).
<sup>b</sup>Subjects with undetermined/missing baseline PCR values were included in the analyses.
<sup>c</sup>Identified criteria are defined by the SMQ terms (narrow scope only) fitting Centers for Disease Control criteria for high-risk for severe SARS-CoV-2 infection.

### Efficacy

#### Incidence of COVID-19

COVID-19 (symptomatic SARS-CoV-2 infection) determination was made by the investigator based on clinical assessment. In a predefined exploratory analysis of efficacy, monthly REGEN-COV reduced the risk of COVID-19 infection over the 6-month dosing interval. There was a 92.4% relative risk reduction (RRR) in COVID-19 infections with REGEN-COV compared to placebo during the treatment period (3/729 [0.4%] vs 13/240 [5.4%]; odds ratio: 0.07 [95% confidence interval (CI), 0.01– 0.27]; nominal *P*-value <0.001) (**Figure 1; Supplementary Table 2**). While not all study participants had completed the follow-up period at the time of data cut-off, during the entire study period there was a 93.0% RRR in COVID-19 infections with REGEN-COV compared to placebo (3/729 [0.4%] vs 14/240 [5.8%]) (**Supplementary Table 2**). These COVID-19 cases were evenly distributed throughout the study (**Supplementary Figure 2**).

**Figure 1.**
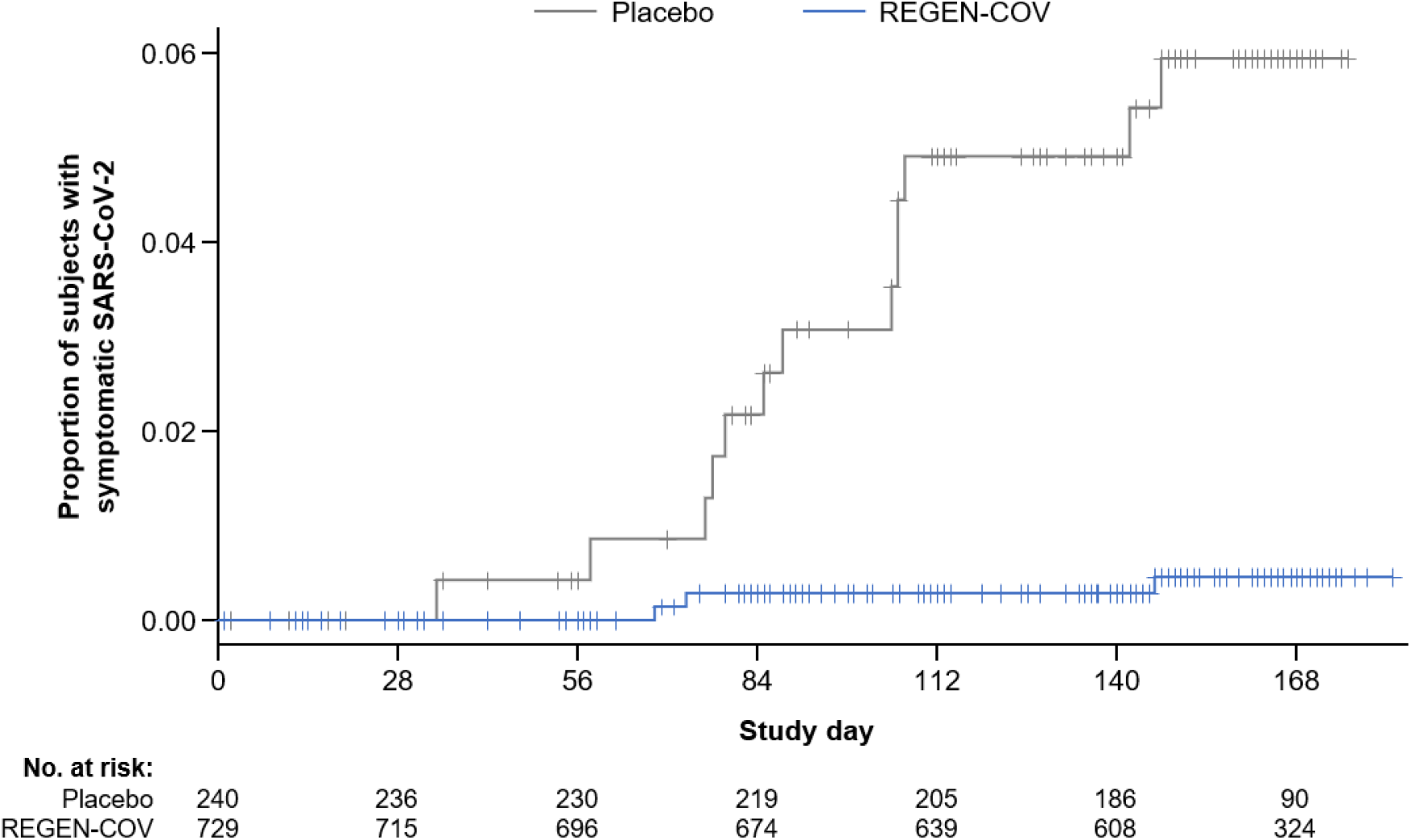
Risk of symptomatic SARS-CoV-2 infection during the treatment period Abbreviations: COVID-19, coronavirus disease 2019; SARS-CoV-2, severe acute respiratory syndrome coronavirus 2. The proportion of subjects with the reported adverse event of SARS-CoV-2 infection compared to the number of subjects at risk for infection in the REGEN-COV group and placebo group is shown here by study day. Symptomatic SARS-CoV-2 (COVID-19) determination was made by the investigator on the basis of clinical assessment.

Two additional subjects who previously received placebo developed COVID-19 infection 2 days and 5 days after COVID-19 vaccination (**Supplementary Table 2; Supplementary Table 3**); these subjects were excluded from the efficacy analysis. In total, including all cases of presumptive COVID-19 regardless of COVID-19 vaccination status, there were 3/729 (0.4%) REGEN-COV and 16/240 (6.7%) placebo recipients who developed COVID-19, corresponding to an approximately 94% RRR, during the study (**Supplementary Table 2; Supplementary Table 3**).

In the REGEN-COV group, each of the 3 (0.4%) subjects who had COVID-19 reported as an AE were seronegative for SARS-CoV-2 antibodies at baseline and at the end-of-treatment period visit (**Supplementary Table 3**). Two of the 3 subjects were negative for SARS-CoV-2 infection as assessed by RT-PCR, and 1 did not have RT-PCR data analyzed. Thus, while counted as a breakthrough from the antibody treatment, none of these 3 patients had laboratory confirmation of infection with SARS-CoV-2. Among these 3 REGEN-COV recipients, symptom duration was 11, 16, and 18 days, and symptoms were mostly mild (**Supplementary Table 3**). In contrast, for the 16 (6.7%) subjects who were reported to have developed COVID-19 in the placebo group, 11 had either seroconversion for SARS-COV-2 anti-nucleocapsid IgG antibodies or a documented SARS CoV-2 infection by RT-PCR or antigen test. Among placebo recipients, symptom duration ranged from 5 to 30 days and symptoms were mostly mild, though a few were reported as moderate in severity. In a sub-group analysis of subjects with laboratory-confirmed SARS-CoV-2 infection, 10/240 (4.2%) in the placebo group and 0/729 in the REGEN-COV group had laboratory-confirmed infection as defined by RT-PCR positivity or seroconversion for SARS-COV-2 anti-nucleocapsid IgG antibodies during the treatment period, corresponding to a 100% RRR for REGEN-COV versus placebo, odds ratio: 0.00 (**Supplementary Table 4**); a listing of all subjects who experienced the AE of COVID-19, including serologic and/or PCR confirmation, is provided in **Supplementary Table 3**.

#### SARS-CoV-2 Seroconversion

Conversion from SARS-CoV-2 anti-nucleocapsid seronegative to seropositive was used to assess the incidence of SARS-CoV-2 infection during the study. Of subjects who were seronegative for anti-spike or anti-nucleocapsid protein (anti-spike immunoglobulin [Ig]G, anti-spike IgA, or anti-nucleocapsid IgG) at baseline, 0/617 (0%) of REGEN-COV recipients were seropositive for anti-nucleocapsid protein (anti-nucleocapsid IgG) compared to 20/208 (9.6%) of placebo recipients at the end-of-treatment period visit, with a 100% risk reduction for anti-SARS-CoV-2 seroconversion, odds ratio: 0.00 (95% CI, 0.00–0.05) (**Supplementary Table 5**). Of the 20 placebo recipients with seroconversion, 8 reported symptomatic SARS-CoV-2 infection during the treatment period (**Supplementary Table 5**).

### Safety and Tolerability

#### Incidence of Treatment-Emergent Adverse Events

Over the 6-month treatment period, there were no AESIs (defined as grade ≥3 ISRs or grade ≥3 hypersensitivity reactions) or deaths reported among any study participants. Serious TEAEs were rare and occurred at similar rates in the REGEN-COV and placebo groups (5/729 [0.7%] vs 2/240 [0.8%]) (**Supplementary Table 6**). No serious TEAEs were assessed as related to study drug. A greater percentage of subjects withdrew from the study due to an AE in the placebo group (12/240 [5.0%]) than in the REGEN-COV group (13/729 [1.8%]) (**Supplementary Table 7**). The most common AE leading to study drug withdrawal was COVID-19 (2/729 [0.3%] REGEN-COV and 11/240 [4.6%] placebo). A slightly higher percentage of subjects reported TEAEs in the REGEN-COV group (400/729 [54.9%]) versus the placebo group (116/240 [48.3%]), which is attributable to the higher rate of ISRs with REGEN-COV (**Table 2**). Aside from ISRs, the only other TEAEs occurring at ≥5% were headache (8.0% vs 7.0%) and COVID-19 (0.4% vs 5.4%) for REGEN-COV versus placebo, respectively (**Supplementary Table 8**).

**Table 2.** Overview of TEAEs in the Treatment Period.

|  | Placebo<br>n = 240 | REGEN-COV<br>1200 mg SC Q4W<br>n = 729 |
| --- | --- | --- |
| Number of TEAEs | 326 | 2107 |
| Number of grade $\geq 3$ TEAEs | 3 | 10 |
| Number of serious TEAEs | 5 | 8 |
| Number of AESIs<sup>a</sup> | 0 | 0 |
| Number of TEAEs resulting in study drug being withdrawn | 13 | 15 |
| Number of TEAEs resulting in death | 0 | 0 |
| Subjects with at least 1 TEAE, n (%) | 116 (48.3) | 400 (54.9) |
| Subjects with at least 1 ISR | 40 (16.7) | 266 (36.5) |
| Grade 1 | 38 (15.8) | 211 (28.9) |
| Grade 2 | 2 (0.8) | 55 (7.5) |
| Grade $\geq 3$ | 0 | 0 |
| Subjects with at least 1 grade $\geq 3$ TEAE<sup>b</sup>, n (%) | 3 (1.3) | 4 (0.5) |
| Subjects with at least 1 serious TEAE, n (%) | 2 (0.8) | 5 (0.7) |
| Subjects with at least 1 AESI, n (%) | 0 | 0 |
| Subjects with at least 1 TEAE resulting in study drug being withdrawn, n (%) | 12 (5.0) | 13 (1.8) |
| Subjects with any TEAE resulting in death, n (%) | 0 | 0 |

Abbreviations: AESI, adverse event of special interest; ISR, injection-site reaction; Q4W, every 4 weeks; SC, subcutaneous; TEAE, treatment emergent adverse event.
<sup>a</sup>AESIs were defined as grade $\geq 3$ injection-site or hypersensitivity reactions.
<sup>b</sup>No grade $\geq 3$ TEAEs were assessed as related to study drug

#### Injection-Site Reactions

In this multi-dose study, ISRs were experienced by 266/729 (36.5%) REGEN-COV recipients and 40/240 (16.7%) placebo recipients during the 6-month treatment period (**Table 2**). All ISRs for REGEN-COV and placebo were mild (grade 1; 28.9% and 15.8%) or moderate (grade 2; 7.5% and 0.8%), respectively (**Table 2**). The majority of ISRs (>60%) resolved within 4 days of dosing (**Supplementary Table 9**), either spontaneously or with the use of over-the-counter treatments. The most prevalent ISRs observed with REGEN-COV were erythema (27.6%) and pruritus (13.0%) (**Supplementary Table 9**).

Across all 7 study sites combined, the rate of REGEN-COV ISRs was stable from dose 1 through 4 (∼13% per dose) and was higher with dose 5 and 6 (∼19% per dose) (**Supplementary Figure 3A**). However, an imbalance in ISR rates was observed between study sites, with the overall incidence being driven by 2 outlier sites (**Supplementary Figure 3B; Supplementary Table 10**); across the other 5 sites, the rate of ISRs was stable across all 6 doses of REGEN-COV (∼6% per dose) (**Supplementary Figure 3C**).

#### Adverse Events Post COVID-19 Vaccination

During this study, 354/969 (36.5%) of all subjects (256/729 [35.1%] REGEN-COV and 98/240 [40.8%] placebo) received COVID-19 vaccination and, per study protocol, were discontinued from further doses of study drug and continued to be followed for safety (**Supplementary Table 11**). Subjects receiving COVID-19 vaccination had a mean of 66.1 (from -20 to 194) days between last dose of study drug and receipt of vaccination, despite Centers for Disease Control and Prevention (CDC) guidelines^24^ recommending a 90-day delay between passive antibody therapy and COVID-19 vaccination (**Supplementary Table 11**). AEs post COVID-19 vaccination occurred in 39 (15.2%) subjects who had previously received REGEN-COV and 18 (18.4%) subjects who had previously received placebo (**Supplementary Table 12**). AEs post COVID-19 vaccination were all grade 1-2, except for a single grade 3 event in a subject previously dosed with REGEN-COV; this grade 3 event of brain mass frontal lobe was assessed as not related to study drug (**Supplementary Table 12**).

### Pharmacokinetics and Immunogenicity

PK analysis showed that concentrations of casirivimab and imdevimab in serum reached steady-state following the third dose of REGEN-COV and were maintained throughout the 6-month treatment period. Concentrations 28 days after the first dose of REGEN-COV were 32.7 mg/L for casirivimab and 24.8 mg/L for imdevimab, with accumulation ratios (ratio of concentrations 28 days after the sixth over the first dose) of 2.08- and 1.98-fold after 6 monthly doses, respectively (**Figure 2**). Notably, in the 3 REGEN-COV recipients who reported COVID-19 symptoms, serum concentration of casirivimab and imdevimab were comparable to subjects without any reported COVID-19 symptoms (**Supplementary Table 13**).

**Figure 2.**
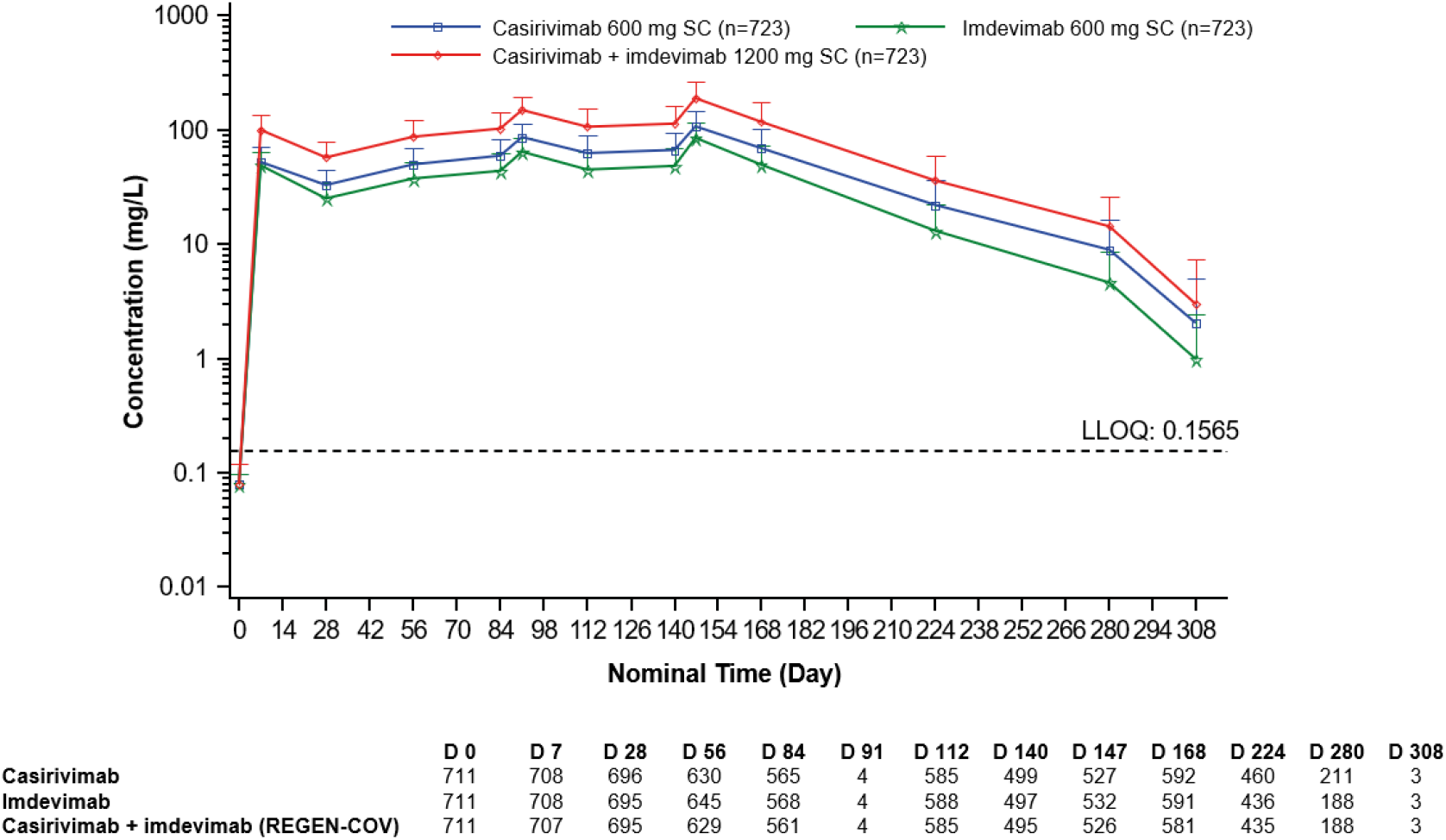
Pharmacokinetics of the concentrations of casirivimab and imdevimab in serum over time Abbreviations: D, day; LLOQ, lower limit of quantification; SC, subcutaneous. Serum concentration of casirivimab, imdevimab, and casirivimab+imdevimab (REGEN-COV) in serum are depicted by study day. Sample collection visits on day 91 and day 208 were included in the original version of the protocol but were removed in protocol amendment 3.

We also examined whether repeat dosing would be associated with the formation of ADAs against either casirivimab or imdevimab, and whether these ADAs would impact circulating levels of drug. Casirivimab and imdevimab both showed low treatment-emergent immunogenicity, measured as ADAs in ≤5% of the population: 1.1% for anti-casirivimab and 5% for anti-imdevimab. Approximately 4% of subjects had pre-existing ADAs against casirivimab (3.1%) and imdevimab (3.9%) (**Table 3**). ADAs did not appear to impact circulating levels of study drug, as concentrations of casirivimab and imdevimab were comparable between subjects with positive and negative ADA results (**Supplementary Figure 4**).

**Table 3.** Casirivimab and Imdevimab Immunogenicity by Anti-Drug Antibody Status.

| ADA Status and Category | Placebo<br>n (%) | REGEN-COV<br>1200 mg SC Q4W<br>n (%) | Overall<br>n (%) |
| --- | --- | --- | --- |
| Casirivimab immunogenicity |  |  |  |
| ADA analysis set | 237 (100) | 717 (100) | 954 (100) |
| Negative | 228 (96.2) | 687 (95.8) | 915 (95.9) |
| Pre-existing immunoreactivity | 8 (3.4) | 22 (3.1) | 30 (3.1) |
| Treatment-boosted response | 0 | 0 | 0 |
| Treatment-emergent response | 1 (0.4) | 8 (1.1) | 9 (0.9) |
| Imdevimab immunogenicity |  |  |  |
| ADA analysis set | 237 (100) | 717 (100) | 954 (100) |
| Negative | 221 (93.2) | 653 (91.1) | 874 (91.6) |
| Pre-existing immunoreactivity | 9 (3.8) | 28 (3.9) | 37 (3.9) |
| Treatment-boosted response | 0 | 0 | 0 |
| Treatment-emergent response | 7 (3.0) | 36 (5.0) | 43 (4.5) |

Abbreviations: AAS, ADA analysis set; ADA, anti-drug antibody; Q4W, every 4 weeks; SC, subcutaneous.
n = number of subjects contributing to each category.

## Discussion

Recent data have shown that some individuals are not able/expected to mount an adequate immune response to COVID-19 vaccination (eg, individuals with immunocompromising conditions, particularly those with B-cell deficiencies, including those taking immunosuppressive medications), leaving them unprotected and at increased risk for SARS-CoV-2 infection and progression to severe COVID-19.^25-27^ Estimates of the number of such immunocompromised individuals are as high as 3% of the US population.^28,29^ This highlights an unmet medical need for a significant subset of the population to have access to a chronic preventive therapy, particularly in light of the ongoing resurgence of COVID-19 cases with the emergence of new variants.

This is the first study to investigate monthly dosing of SC administered REGEN-COV. Results from this study show that REGEN-COV is well-tolerated, and extend the findings of the household contact prevention study^17^ by showing that monthly REGEN-COV prevents COVID-19 over a 6-month period. In a predefined exploratory efficacy analysis, REGEN-COV treatment resulted in a 92.4% RRR in presumptive COVID-19 versus placebo and a 100% RRR in laboratory-confirmed COVID-19. Although an assessment of efficacy was not the primary purpose of the study, the RRR for the development of COVID-19 seen in subjects treated with REGEN-COV is striking, and similar to the risk reduction seen in the vaccine trials.^30-33^

Results of this study also demonstrate a significant reduction in anti-nucleocapsid IgG seroconversion among REGEN-COV treated subjects over the 6-month course of therapy. The production of SARS-CoV-2 anti-nucleocapsid IgG antibodies, as assessed by seroconversion from a seronegative status at baseline to a seropositive status, was considered a proxy of SARS-CoV-2 infection. There was a 9.6% seroconversion rate among placebo recipients compared to the absence of seroconversion (0%) among those who received REGEN-COV, suggesting that REGEN-COV is also effective in preventing asymptomatic SARS-CoV-2 infection when given as chronic treatment for pre-exposure prophylaxis of COVID-19. This finding is particularly impactful for immunocompromised individuals and in the context of long-term care facilities, such as nursing homes, where protection against asymptomatic infection could reduce viral transmission and potentially decrease overall morbidity and mortality in these vulnerable populations.^34-36^

Multiple dose administration of REGEN-COV was well-tolerated. Compared to placebo, REGEN-COV treatment was associated with a slightly higher frequency of ISRs. The overall rate of ISRs with REGEN-COV was ∼13% per dose, and 36.5% across all 6 doses combined. However, there was significant variability in the ISR rate between sites, with 2 outlier sites driving the incidence. A previous phase 3 study assessing the efficacy of REGEN-COV 1200 mg as post-exposure prophylaxis has shown an ISR rate of 4% after a single SC dose.^17^ This is consistent with the reported ISR rate observed at 5 of the 7 sites in this study (∼6% per dose); however, 2 sites exhibited disproportionately higher ISR rates of 19-34% and 17-54% per dose, respectively. All ISRs were mild to moderate in severity (grade 1–2) and the incidence and severity of ISRs did not substantially increase with repeat dosing. There were no grade ≥3 hypersensitivity reactions in this study, and repeated dosing of REGEN-COV showed low treatment-emergent immunogenicity. Concentrations of each component of REGEN-COV, casirimivab and imedivimab, were maintained in the projected therapeutic range for viral neutralization throughout the dosing interval.

A potential limitation of this study was that regular RT-PCR testing was not performed in all participants who presented with a clinical syndrome compatible with COVID-19. However, when only considering laboratory confirmed SARS-CoV-2 infection, REGEN-COV showed a 100% RRR in the development of COVID-19. The discrepancy could imply that these subjects had illnesses that were not due to SARS-CoV-2, or that sample collection was not close enough to symptom onset.

An additional limitation of this study is that it was conducted before the emergence of several SARS-CoV-2 variants. However, extensive non-clinical testing has demonstrated that REGEN-COV retains its neutralization capacity against all clinically relevant viral variants tested to date, including beta, gamma, epsilon, and delta variants.^10,13^

Although it should not be deemed an appropriate substitute for vaccination in immunocompetent individuals, the efficacy and safety profile of REGEN-COV demonstrated in this study strongly support that REGEN-COV should be used for chronic prevention of COVID-19 in individuals not expected to mount a sufficient immune response to vaccination.

## Supporting information

Author COI

Author COI

Author COI

Author COI

Author COI

Author COI

Author COI

Author COI

Author COI

Author COI

Author COI

Author COI

Author COI

Author COI

Author COI

Author COI

Author COI

Author COI

Author COI

Author COI

Author COI

Author COI

Author COI

Author COI

Author COI

Author COI

Author COI

Author COI

Author COI

Author COI

## Data Availability

Qualified researchers may request access to study documents (including the clinical study report, study protocol with any amendments, blank case report form, and statistical analysis plan) that support the methods and findings reported in this manuscript. Individual anonymized participant data will be considered for sharing once the indication has been approved by a regulatory body, if there is legal authority to share the data and there is not a reasonable likelihood of participant re-identification. Submit requests to https://vivli.org/.

## Acknowledgments

The authors would like to thank the participants who volunteered for the study; the investigators involved in this study; Kaitlyn Scacalossi, PhD, and Caryn Trbovic, PhD, from Regeneron Pharmaceuticals for medical writing support; and Prime, Knutsford, UK, for formatting and copy-editing suggestions.

