## Supplemental Materials for "Repeat Subcutaneous Administration of REGEN-COV^®^ in Adults is Well-Tolerated and Prevents the Occurrence of COVID-19"

SUPPLEMENTARY APPENDIX

Study Sites and Investigators

**Celerion, Tempe, Arizona:** Danielle Armas, Mark Allison, Mayra Alvarado, Nicole Bartreau, Kelly Black, John Laabs, Terry O'Reilly, Jill Patterson, Jeanmarie Ricks, Trisha Tavares, Tracy Vincent, Weston Young

**Celerion, Lincoln, Nebraska:** Scott Rasmussen, Samantha Boardman, Brent Bunz, Allen Hunt, Lisa Mapson, Miri Pelc, Robert Schwab, Natasha Wilson

**Benchmark Research, Sacramento, California:** Masaru Oshita, Lisa Anderson, Kim Blossom, Michael Cancilla, Shannon Chand, Angella Cuellar, Yaman Darmarathne, Shaila Faulkner, Yana Gordeyeva, Lizban Habte, Greg Hachigian, Michelle Hisey, Richard Joven, Zuriel Joven, Ashely Jungsten

**Central Texas Research, Austin, Texas:** Cynthia Brinson, David Wright

**Midwest Clinical Research, Dayton, Ohio:** Steven Folkerth, Otto Dueno, Isabel Kuhar-Arcure, Christine Dyson, Melissa Ellis, Lauren Hoke, Scott Kustowski, Nita Miller, Rebecca Vernon

**Woodland Research Northwest, Rogers, Arkansas:** Shari De Silva, Robert Billingsley

**Clinical Pharmacology of Miami, Miami, Florida:** Juan-Carlos Rondon, Juan Alvarez, Joan Beltran, Ernesto Fuentes, Kenneth Lasseter, Samuel Oberstein, Charlene Soler, Pablo Suso, Jorge Urgell

### Regeneron Study Team

Adriel Perez, Andrea Hooper, Bala Dass, Bobbi Zastrie, Brian Head, Bryan Zhu, Carol Lee, Caryn Trbovic, Cathrine Ogunjobi, Claxton Whiteside, Debbie Bochenek, Dipinder Kaur, Dona Bianco, Donna Cohen, Eduardo Forleo-Neto, Elisa Halker, Evelyn Gasparino, Flonza Isa, George Vlamis, Ingeborg Heirman, Jaclyn Patterson, Jennifer Hamilton, Jessica Border, John Rembis, Jordan Ursino, Jonathan Meyer, Joseph Wolken, Kaitlyn Scacalossi, Kalpana Pullakhandam, Karen Thabet, Kenneth Turner, Kurt Ringleben, Lanesha Hill, Lisa Sherpinsky, Lori Faria, Marc Dickens, Meagan O’Brien, Michael Klinger, Michael Partridge, Nitin Kumar, Qin Li, Rachael Webster, Sabrina Post, Samit Ganguly, Shikha Bansal, Somang Kim, Soraya Nossoughi, Steven Elkin, Susan Irvin, Susan Olobashola, Teresa Rodriguez, Viral Seth, Wenjun Zheng, Yanmei Tian, Yunji Kim, Zachary Kendall

### SUPPLEMENTARY METHODS

#### Study Oversight

The protocol was developed by the sponsor (Regeneron Pharmaceuticals, Inc.). Data were collected by the study investigators and analyzed by the sponsor. The trial was conducted in accordance with the principles of the Declaration of Helsinki, International Council for Harmonisation Good Clinical Practice guidelines, and applicable regulatory requirements. All patients provided written informed consent before participating in the trial. Ethics approval was obtained from the Western Institutional Review Board, Puyallup, WA, USA.

#### Dose Rationale

The administration of the 1200 mg subcutaneous (SC) dose was based on the goal of exceeding a target concentration of 20 μg/mL for each monoclonal antibody in serum for 28 days after dosing in ≥95% of subjects. By achieving this target concentration in serum, concentrations of each monoclonal antibody in lung epithelial lining fluid were anticipated to meet or exceed those required for 99% neutralization of severe acute respiratory syndrome coronavirus 2 (SARS-CoV-2) *in vitro*.

#### Dose Administration

Each dose was given as 4 SC injections of 2.5 mL (120 mg/mL), with each injection administered to 1 of 6 different anatomical locations (4 abdominal quadrants and both thighs). Randomization was stratified by age ≥50 years and presence or absence of any of the following chronic medical conditions: cardiovascular disease (including hypertension); chronic lung disease (including asthma); chronic metabolic disease (including diabetes); chronic kidney disease, or chronic liver disease.

#### Inclusion and Exclusion Criteria

##### Inclusion Criteria

A subject must meet the following criteria to be eligible for inclusion in the study:

1. 18 to 90 years of age (inclusive) at the signing of informed consent
2. Healthy or has chronic medical condition(s) that is stable and well controlled as per the opinion of the investigator and is not likely to require medical intervention through the end of study
3. Stable medication for co-morbid condition(s) for at least 6 months prior to screening
4. Willing and able to comply with study visits and study-related procedures, including compliance with site precautionary requirements related to SARS-CoV-2 infection and transmission
5. Willing and able to provide signed informed consent

##### Exclusion Criteria

A subject who meets any of the following criteria will be excluded from the study:

1. Positive diagnostic test for SARS-CoV-2 infection ≤72 hours prior to randomization

*Note: This test is done as part of screening. The sample for the test should be collected ≤72 hours within randomization, and the result should be reviewed and confirmed negative prior to dosing.*

2. Subject-reported clinical history of coronavirus disease 2019 (COVID-19) as determined by investigator
3. Subject-reported history of prior positive diagnostic test for SARS-CoV-2 infection
4. Active respiratory or non-respiratory symptoms suggestive or consistent with COVID‑19
5. Medically attended acute illness, systemic antibiotics use, or hospitalization (ie, >24 hours) for any reason within 30 days prior to screening
6. Clinically significant abnormal laboratory results at screening as defined by 1 or more of the following (may be repeated once):

- Glycated hemoglobin (HbA1c) ≥8.0%
- Hemoglobin <10 g/dL
- Absolute neutrophil count <1.5 x 10^9^/L
- Platelet count <75 x 10^9^/L
- Serum creatinine >1.5x upper limit of normal (ULN) or estimated glomerular filtration rate ≤60 mL/min/1.73m^2^
- Hepatic function abnormalities defined as 1 or more of the following:
  - Aspartate aminotransferase (AST) and/or alanine aminotransferase (ALT), and/or alkaline phosphatase (ALP) >2x ULN
  - Total bilirubin >1x ULN

7. Acute exacerbation of a chronic pulmonary condition (eg, chronic obstructive pulmonary disease, asthma exacerbations) in the past 6 months prior to screening
8. Abnormal blood pressure (BP) at screening, as defined by diastolic BP >100 mm Hg and/or systolic BP >160 mm Hg. BP measurements may be repeated once at screening
9. History of heart failure hospitalization, diagnosis of a myocardial infarction, stroke, transient ischemic attack, unstable angina, percutaneous or surgical revascularization procedure (coronary, carotid, or peripheral vascular), or intracardiac device placement (eg, pacemaker) within 12 months prior to screening
10. Cancer requiring treatment currently or in the past 5 years, except for non-melanoma skin cancer or cervical/anus cancer in-situ
11. History of significant multiple and/or severe allergies (eg, latex gloves), or has had an anaphylactic reaction to prescription or non-prescription drugs or food. This is to avoid potential confounding of the safety data and not due to a particular safety risk.
12. Treatment with another investigational drug in the last 30 days or within 5 half-lives of the investigational drug, whichever is longer, prior to screening
13. Received investigational or approved SARS-CoV-2 vaccine
14. Received investigational or approved passive antibodies for SARS-CoV-2 infection prophylaxis (eg, convalescent plasma or sera, monoclonal antibodies, hyperimmune globulin)
15. Use of remdesivir, intravenous immunoglobulin (IVIG), or other anti-SARS viral agents within 2 months prior to screening
16. Regular alcohol consumption of ≥21 drinks per week
17. Member of the clinical site study team and/or immediate family
18. Pregnant or breastfeeding women
19. Women of childbearing potential^a^ who are unwilling to practice highly effective contraception prior to the initial dose/start of the first treatment, during the study, and for at least 8 months after the last dose. Highly effective contraceptive measures include:
    a. Stable use of combined (estrogen- and progestogen-containing) hormonal contraception (oral, intravaginal, transdermal) or progestogen-only hormonal contraception (oral, injectable, implantable) associated with inhibition of ovulation initiated 2 or more menstrual cycles prior to screening
    b. Intrauterine device (IUD) or intrauterine hormone-releasing system (IUS)
    c. Bilateral tubal ligation
20. Sexually active men who are unwilling to use the following forms of medically acceptable birth control during the study drug follow-up period and for 8 months after the last dose of study drug: vasectomy with medical assessment of surgical success OR consistent use of a condom. Sperm donation is prohibited during the study and for up to 8 months after the last dose of study drug.

^a^Women of childbearing potential are defined as women who are fertile following menarche until becoming postmenopausal, unless permanently sterile. Permanent sterilization methods include hysterectomy, bilateral salpingectomy, and bilateral oophorectomy. A postmenopausal state is defined as no menses for 12 months without an alternative medical cause. A high follicle stimulating hormone (FSH) level in the postmenopausal range may be used to confirm a postmenopausal state in women not using hormonal contraception or hormonal replacement therapy. However, in the absence of 12 months of amenorrhea, a single FSH measurement is insufficient to determine the occurrence of a postmenopausal state. The above definitions are according to the Clinical Trial Facilitation Group guidance. Pregnancy testing and contraception are not required for women with documented hysterectomy or tubal ligation.

#### Diagnosis of COVID-19 During the Study

Subjects were provided instructions to promptly notify site personnel by phone with any changes in their health status. In cases of suspected COVID-19, subjects were to undergo an unscheduled visit for evaluation of symptoms and to assess for SARS-CoV-2 infection. The assessment of SARS-CoV-2 infection may have been performed by central laboratory reverse transcription polymerase chain reaction (RT-PCR) or by an approved or authorized diagnostic assay performed according to the site’s standards and procedures. Alternatively, subjects were permitted to provide documentation of a positive test in lieu of a confirmatory test, if allowable by site standards and procedures. Both clinically defined COVID-19 cases (as determined by the investigator) and laboratory-confirmed COVID-19 cases (with documentation from an approved test) cases were reported as adverse events (AEs) during the study

#### Assessment of SARS-CoV-2 Serostatus

To assess immunity to SARS-CoV-2, SARS-CoV-2 antibodies were characterized at baseline by assays for anti-S1 domain of spike protein immunoglobulin (Ig)G and IgA antibodies (EuroImmun Anti-SARS-CoV-2 enzyme-linked immunosorbent assay [ELISA]) and anti-nucleocapsid IgG antibodies (Abbott SARS-Cov-2 IgG Architect). At the end of the treatment and follow up periods, SARS-CoV-2 antibodies were only assessed by the anti-nucleocapsid IgG (Abbott) assay, as anti-nucleocapsid IgG is considered indicative of infection (rather than vaccination) and REGEN-COV does not interfere with this assay. The presence or absence of anti-spike antibodies was not assessed post baseline as REGEN-COV may interfere with anti-spike serologic assays.

#### COVID-19 Vaccination During the Study

COVID-19 vaccination became available in the US starting in December 2020, several months after this trial began on July 26, 2020. Subjects enrolled in this study who elected to receive COVID-19 vaccination were discontinued from study drug and entered follow-up. COVID-19 vaccination tolerability was assessed by AE reporting following each dose of the vaccine; only unsolicited AEs were captured. COVID-19 vaccines received by subjects were based on vaccine availability and subject selection.

The Centers for Disease Control and Prevention guidelines suggested a 90-day waiting period between passive antibody therapy (such as REGEN-COV) and COVID-19 vaccination (<https://www.cdc.gov/vaccines/covid-19/clinical-considerations/covid-19-vaccines-us.html>); however, as these recommendations were made specifically for patients receiving antibody therapy for the treatment of active COVID-19, this guidance was not always followed by subjects and/or investigators, with some subjects receiving COVID-19 vaccination <90 days from their last dose of study drug. AEs following COVID-19 vaccination are presented herein. A separate sub-study examining the potential of REGEN-COV to impact the immune response following COVID-19 vaccination is ongoing.

#### Pharmacokinetic Analysis Methods

The pharmacokinetic (PK) analysis population included all participants who received any study drug (safety population) and who had ≥1 non-missing result following the first dose of study drug. Participants were analyzed based on actual treatment received. Overall, the PK analysis set was comprised of 723 subjects. Blood samples for measurement of casirivimab and imdevimab concentrations in serum were collected from participants randomized to 1200 mg SC or placebo at predose and on days 1, 8, 29, 57, 85, 113, 141, 148, 169, 225, 281, and 365, and the end-of-treatment visit. Serum samples were analyzed for concentrations of total casirivimab and total imdevimab using a validated electrochemiluminescence immunoassay with a lower limit of quantification of 0.156 mg/L. From the concentration in serum data at the above time points, descriptive analysis of pharmacokinetics of casirivimab and imdevimab were performed.

#### Statistical Analysis

##### Handling of Missing Data

Missing data for summary analyses were handled as follows: if the severity of a TEAE was missing, it was classified as “severe” or “grade 3.” When the partial AE date/time information did not indicate that the AE started prior to or after study treatment, the AE was classified as treatment-emergent. If a subject had a missing baseline value, the subject was grouped in the category “normal/missing at baseline,” where baseline was defined as the last assessment obtained before the first dose of study drug.

Supplementary Table 1. Subjects with Medical History Identified as High-Risk for Severe SARS-CoV-2 Infection

|  | Placebo n = 240 | REGEN-COV 1200 mg SC Q4W n = 729 | Total N = 969 |
| --- | --- | --- | --- |
| Subjects with at least 1 medical history condition identified as high-risk for severe SARS-CoV-2 infection^a^, n (%) | 42 (17.5) | 152 (20.9) | 194 (20.0) |
| Medical history category, n (%) | | | |
| Hypertension | 29 (12.1) | 113 (15.5) | 142 (14.7) |
| Asthma | 14 (5.8) | 36 (4.9) | 50 (5.2) |
| Asthma exercise induced | 1 (0.4) | 1 (0.1) | 2 (0.2) |
| Blood pressure increased | 1 (0.4) | 1 (0.1) | 2 (0.2) |
| Coronary artery bypass | 1 (0.4) | 3 (0.4) | 4 (0.4) |
| Coronary artery disease | 1 (0.4) | 7 (1.0) | 8 (0.8) |
| Coronary artery occlusion | 1 (0.4) | 0 | 1 (0.1) |
| Myocardial infarction | 1 (0.4) | 4 (0.5) | 5 (0.5) |
| Angina pectoris | 0 | 1 (0.1) | 1 (0.1) |
| Bronchial hyperreactivity | 0 | 2 (0.3) | 2 (0.2) |
| Coronary arterial stent insertion | 0 | 3 (0.4) | 3 (0.3) |
| Pre-eclampsia | 0 | 1 (0.1) | 1 (0.1) |
| Renal failure | 0 | 2 (0.3) | 2 (0.2) |

Abbreviations: BMI, body mass index; Q, quartile; Q4W, every 4 weeks; SC, subcutaneous; SD, standard deviation; SMQ, standardized Medical Dictionary of Regulatory Activities queries.

^a^Identified criteria are defined by the SMQ terms (narrow scope only) of hypertension, ischemic heart disease, asthma/bronchospasm, interstitial lung disease, and chronic kidney disease; and high-level group terms of arteriosclerosis, stenosis, vascular insufficiency and necrosis and glucose metabolism disorders (including diabetes mellitus); and high level terms of transient cerebrovascular events, bronchospasm and obstruction, lower respiratory tract inflammatory and immunologic conditions, hepatocellular damage and hepatitis NEC, hepatic vascular disorders, hepatic failure and associated disorders, and hepatic fibrosis and cirrhosis; and preferred terms of cerebrovascular accident and emphysema.

Supplementary Table 2. Proportion of Subjects with Symptomatic SARS-CoV-2 Infection or Seroconversion

|  | Placebo (n = 240) | REGEN-COV 1200 mg SC Q4W (n = 729) |
| --- | --- | --- |
| Subjects with symptomatic SARS-CoV-2 infections^a^ during the treatment period, n (%) | 13 (5.4) | 3 (0.4) |
| Risk reduction of symptomatic SARS-CoV-2 infections, % |  | 92.4 |
| Odds ratio estimate (drug vs placebo) |  | 0.07 |
| 95% CI |  | 0.01–0.27 |
| *P*-value versus placebo^b^ |  | <.0001 |
| Subjects with symptomatic SARS-CoV-2 infections^a^ during the entire study period, n (%) | 14 (5.8) | 3 (0.4) |
| Risk reduction of symptomatic SARS-CoV-2 infections, % |  | 93.0 |
| Odds ratio estimate (drug vs placebo) |  | 0.07 |
| 95% CI |  | 0.01–0.24 |
| *P*-value versus placebo^b^ |  | <.0001 |

Abbreviations: CI, confidence interval; COVID-19, coronavirus disease 2019; Q4W, every 4 weeks; RT-PCR, reverse transcription polymerase chain reaction; SARS-CoV-2, severe acute respiratory syndrome coronavirus 2; SC, subcutaneous.

^a^Symptomatic SARS-CoV-2 infection was defined clinically by the investigator and reported as an adverse event; RT-PCR testing was not required. Included are events that were reported before the COVID-19 vaccination date, if any. Not included in this table: 2 subjects in the placebo group developed symptomatic SARS-CoV-2 infection after vaccination and are not included in the treatment period or entire study period; these subjects continued to be followed for safety.

_b_Nominal *P*-value from Fisher’s exact test is provided.

Supplementary Table 3. Listing of Subjects with Symptomatic COVID-19 Infection During the Treatment Period

| Age | Sex | Medical History | Study Arm | Study Day of Start of COVID-19 Event | Duration of Event (Days) | Baseline Serology Results | | Serology & PCR Confirmation of COVID-19 | | | | |
| --- | --- | --- | --- | --- | --- | --- | --- | --- | --- | --- | --- | --- |
|  |  |  |  |  |  |  |  | End-of-Treatment Period Visit Serology Results | | Central Lab PCR | | Local Lab Test |
|  |  |  |  |  |  | Baseline Serology Status | Study Day of Baseline Serology | End-of-Treatment Period Visit Serology Status (IgG Anti‑N) | Study Day of End-of-Treatment Period Visit Serology  (Days After Onset of COVID-19 Event) | Central Lab PCR | Study Day of Central Lab PCR  (Days After Onset of COVID-19 Event) | Local Lab Test & Result (Study Day of Local Lab) |
| Symptomatic COVID-19 cases that occurred in the treatment period | | | | | | | | | | | | |
| 21–25 | M | None | REGEN-COV 1200 mg | 146 | 11 | - | 1 | - | 171 (25) | - | 157 (11) |  |
| 21–25 | M | None | REGEN-COV 1200 mg | 68 | 16 | - | 1 | - | 85 (17) | ND |  |  |
| 56–60 | F | Cholecystectomy, cholelithiasis, depression, gastric bypass, headache, hysterectomy, insomnia, heavy menstrual bleeding, migraine, obesity, restless legs syndrome | REGEN-COV 1200 mg | 73 | 18 | - | 1 | - | 93 (20) | - | 79 (6) | PCR- (79) |
| 66–70 | F | Abdominal hernia, female sterilization, abdominoplasty | Placebo | 34 | 30 | - | 1 | + | 56 (22) | + | 37 (3) |  |
| 61–65 | F | Mammoplasty, varicose vein operation, seasonal allergy, female sterilization, abdominoplasty, varicose vein | Placebo | 79 | 5 | - | 1 | + | 91 (12) | + | 83 (4) |  |
| 41–45 | F | Caesarean section | Placebo | 147 | 94 | - | 1 | + | 171 (24) | + | 151 (4) |  |
| 26–30 | F | Anxiety, myringotomy, cholecystectomy, cholelithiasis, ear infection, drug hypersensitivity, hypercholesterolemia, staphylococcal skin infection, hand fracture, fracture treatment | Placebo | 77 | 29 | - | 1 | - | 91 (14) | + | 84 (7) | PCR+ (84) |
| 31–35 | F | Acne, rhinoplasty, female sterilization | Placebo | 105 | 7 | - | 1 | - | 116 (11) | + | 106 (1) |  |
| 41–45 | M | Appendicectomy, appendicitis, pneumothorax, amblyopia, physiotherapy, pneumonia, pneumonia, amblyopia therapy | Placebo | 88 | 11 | - | 1 | + | 113 (25) | ND |  | PCR+ (91) |
| 41–45 | M | Eczema, epididymectomy, epididymitis, seasonal allergy, vasectomy | Placebo | 106 | 13 | - | 1 | + | 130 (24) | ND |  |  |
| 61–65 | M | Deep vein thrombosis, Factor V Leiden mutation, hyperlipidemia, sleep apnea syndrome, pulmonary embolism, pyloric stenosis, pyloromyotomy, fracture treatment, vasectomy | Placebo | 85 | 14 | - | 1 | + | 111 (26) | ND |  |  |
| 31–35 | F | Cervical dysplasia, seasonal allergy, loop electrosurgical excision procedure | Placebo | 106 | 10 | - | 1 | + | 128 (22) | ND |  |  |
| 41–45 | M | HIV infection, hyperlipidemia | Placebo | 107 | 13 | - | 1 | + | 126 (19) | ND |  |  |
| 46–50 | M | Anxiety, small intestine carcinoma, colectomy, post-traumatic stress disorder, dermal cyst, sleep apnea syndrome | Placebo | 76 | 8 | - | 1 | ND |  | ND |  | Ag+ (N/A) |
| 46–50 | F | Pain, hysterectomy, insomnia | Placebo | 58 | 11 | - | 1 | ND |  | ND |  |  |
| 56–60 | M | Hypercholesterolemia, tonsillitis | Placebo | 142 | 9 | - | 1 | Withdrew consent |  | ND |  |  |
| Symptomatic COVID-19 cases that were not in the treatment period | | | | | | | | | | | | |
| 31–35^a^ | M | Gastroesophageal reflux disease, back pain | Placebo | 137 | 7 | - | 1 | - |  | ND |  |  |
| 56–60^b^ | M | Hypertension | Placebo | 125 | 15 | + | 1 | + | 141 (16) | ND |  |  |
| 26–30^c^ | M | Drug hypersensitivity, mammoplasty | Placebo | 57 | 11 | + | 1 | ND |  | ND |  |  |

Abbreviations: COVID-19, coronavirus disease 2019; Ig, immunoglobulin; PCR, polymerase chain reaction; ND, no data.

^a^Subject had a COVID-19 event after receiving COVID-19 vaccination on study day 132.

^b^Subject had a COVID-19 event after receiving COVID-19 vaccination on study day 122.

^c^Subject had a COVID-19 event while in follow-up.

Supplementary Table 4. Proportion of Subjects Who Had a Confirmed Symptomatic SARS-CoV-2 Infection During the Treatment Period

|  | Placebo  (n = 240) | REGEN-COV 1200 mg SC Q4W  (n = 729) |
| --- | --- | --- |
| Subjects with symptomatic SARS-CoV-2 infections, n (%) | 10 (4.2) | 0 |
| Risk reduction of symptomatic SARS-CoV-2 infections, % |  | 100 |
| Odds ratio estimate (drug vs placebo) |  | 0.00 |

Abbreviations: COVID-19, coronavirus disease 2019; Q4W, every 4 weeks; RT-PCR, reverse transcription polymerase chain reaction; SARS-CoV-2, severe acute respiratory syndrome coronavirus 2; SC, subcutaneous.

Symptomatic SARS CoV-2 infection was defined clinically by the investigator and reported as an adverse event. Only the symptomatic SARS CoV-2 infections confirmed with serology testing or central RT-PCR testing were included. Included are events that were reported before the COVID-19 vaccination date, if any.

Supplementary Table 5. Proportion of Subjects with SARS-CoV-2 Seroconversion

|  | Placebo (n = 240) | REGEN-COV 1200 mg SC Q4W (n = 729) |
| --- | --- | --- |
| Subjects with anti SARS-CoV-2 negative serology test results at baseline, n^a^ | 208 | 617 |
| Subjects with anti SARS-CoV-2 anti-nucleocapsid antibody seroconversion at end-of-treatment period visit, n (%)^b.c^ | 20 (9.6) | 0 |
| Risk reduction of anti SARS-CoV-2 serology test, % |  | 100 |
| Odds ratio estimate (drug vs placebo) |  | 0.00 |
| 95% CI |  | (0.00 to 0.05) |

Abbreviations: CI, confidence interval; Ig, immunoglobulin; Q4W, every 4 weeks; SARS-CoV-2, severe acute respiratory syndrome coronavirus 2; SC, subcutaneous.

^a^Baseline serology characterized anti SARS-CoV-2 anti-N and anti-S protein antibodies (anti-S1 domain of spike protein IgG and IgA antibodies [EuroImmun] and anti-nucleocapsid IgG antibodies [Abbott]); negative at baseline means negative for all, positive at baseline means positive for any.

^b^End-of-treatment/follow-up period serology characterized anti-N protein antibodies (anti-nucleocapsid IgG [Abbott]).

^c^Seroconversion is defined as seronegative serology at baseline and seropositive serology at the end-of-treatment period visit.

Supplementary Table 6. Subjects with Serious TEAEs in the Treatment Period

| Primary System Organ Class, Preferred Term, n (%) | Placebo (n = 240) | REGEN-COV 1200 mg SC Q4W  (n = 729) |
| --- | --- | --- |
| Subjects with at least 1 serious TEAE | 2 (0.8) | 5 (0.7) |
| Cardiac disorders | 0 | 1 (0.1) |
| Angina pectoris | 0 | 1 (0.1) |
| Gastrointestinal disorders | 1 (0.4) | 1 (0.1) |
| Colitis | 0 | 1 (0.1) |
| Enteritis | 1 (0.4) | 0 |
| Injury,­ poisoning and procedural complications | 0 | 1 (0.1) |
| Procedural pain | 0 | 1 (0.1) |
| Musculoskeletal and connective tissue disorders | 1 (0.4) | 1 (0.1) |
| Spinal instability | 0 | 1 (0.1) |
| Pain in extremity | 1 (0.4) | 0 |
| Nervous system disorders | 1 (0.4) | 1 (0.1) |
| Transient ischaemic attack | 0 | 1 (0.1) |
| Thoracic outlet syndrome | 1 (0.4) | 0 |
| Psychiatric disorders | 0 | 1 (0.1) |
| Major depression | 0 | 1 (0.1) |
| Post-traumatic stress disorder | 0 | 1 (0.1) |
| Vascular disorders | 1 (0.4) | 1 (0.1) |
| Aortic aneurysm | 0 | 1 (0.1) |
| Subclavian vein thrombosis | 1 (0.4) | 0 |
| General disorders and administration site conditions | 1 (0.4) | 0 |
| Peripheral swelling | 1 (0.4) | 0 |

Abbreviations: COVID-19, coronavirus disease 2019; Q4W, every 4 weeks; SC, subcutaneous; TEAE, treatment-emergent adverse event.

Medical Dictionary of Regulatory Activities (Version 24.0) coding dictionary applied. Included are events that were reported before the COVID-19 vaccination date, if any. No serious TEAEs were assessed as related to study drug in either treatment group.

Supplementary Table 7. TEAEs Leading to Study Drug Being Withdrawn in the Treatment Period

| Primary System Organ Class,  Preferred Term, n (%) | Placebo (n = 240) | REGEN-COV 1200 mg SC Q4W  (n = 729) |
| --- | --- | --- |
| Subjects with at least 1 TEAE resulting in study drug being withdrawn | 12 (5.0) | 13 (1.8) |
| Infections and infestations | 11 (4.6) | 4 (0.5) |
| COVID-19 | 11 (4.6) | 2 (0.3) |
| Asymptomatic COVID-19 | 0 | 1 (0.1) |
| Viral infection | 0 | 1 (0.1) |
| Skin and subcutaneous tissue disorders | 0 | 3 (0.4) |
| Alopecia | 0 | 1 (0.1) |
| Pruritus | 0 | 1 (0.1) |
| Urticaria | 0 | 1 (0.1) |
| Blood and lymphatic system disorders | 1 (0.4) | 1 (0.1) |
| Lymphadenopathy | 0 | 1 (0.1) |
| Anemia | 1 (0.4) | 0 |
| Cardiac disorders | 0 | 1 (0.1) |
| Angina pectoris | 0 | 1 (0.1) |
| Gastrointestinal disorders | 0 | 1 (0.1) |
| Abdominal pain | 0 | 1 (0.1) |
| Investigations | 0 | 1 (0.1) |
| Blood creatine phosphokinase increased | 0 | 1 (0.1) |
| Nervous system disorders | 0 | 1 (0.1) |
| Transient ischemic attack | 0 | 1 (0.1) |
| Psychiatric disorders | 0 | 1 (0.1) |
| Major depression | 0 | 1 (0.1) |
| Post-traumatic stress disorder | 0 | 1 (0.1) |
| Vascular disorders | 0 | 1 (0.1) |
| Aortic aneurysm | 0 | 1 (0.1) |
| Metabolism and nutrition disorders | 1 (0.4) | 0 |
| Fluid retention | 1 (0.4) | 0 |

Abbreviations: COVID-19, coronavirus disease 2019; Q4W, every 4 weeks; SC, subcutaneous; TEAE, treatment-emergent adverse event.

Medical Dictionary of Regulatory Activities (Version 24.0) coding dictionary applied. Included are events that were reported before the COVID-19 vaccination date, if any.

Supplementary Table 8. TEAEs by Preferred Term (≥1% Occurrence in Any Treatment Group) in the Treatment Period

| Primary System Organ Class Preferred Term, n (%) | Placebo (n = 240) | REGEN-COV 1200 mg SC Q4W (n = 729) |
| --- | --- | --- |
| Subjects with at least 1 TEAE | 116 (48.3) | 400 (54.9) |
| General disorders and administration-site conditions | 48 (20.0) | 287 (39.4) |
| Injection-site reaction | 40 (16.7) | 266 (36.5) |
| Fatigue | 6 (2.5) | 20 (2.7) |
| Chills | 2 (0.8) | 8 (1.1) |
| Pain | 4 (1.7) | 7 (1.0) |
| Nervous system disorders | 27 (11.3) | 85 (11.7) |
| Headache | 17 (7.1) | 58 (8.0) |
| Dizziness | 3 (1.3) | 11 (1.5) |
| Migraine | 3 (1.3) | 3 (0.4) |
| Gastrointestinal disorders | 20 (8.3) | 72 (9.9) |
| Nausea | 6 (2.5) | 24 (3.3%) |
| Diarrhea | 4 (1.7) | 14 (1.9) |
| Abdominal pain | 0 | 10 (1.4) |
| Vomiting | 0 | 7 (1.0) |
| Respiratory,­ thoracic, and mediastinal disorders | 16 (6.7) | 50 (6.9) |
| Oropharyngeal pain | 6 (2.5) | 18 (2.5) |
| Nasal congestion | 2 (0.8) | 14 (1.9) |
| Cough | 2 (0.8) | 13 (1.8) |
| Rhinorrhea | 3 (1.3) | 10 (1.4) |
| Infections and infestations | 28 (11.7) | 51 (7.0) |
| Upper respiratory tract infection | 3 (1.3) | 11 (1.5) |
| Viral infection | 3 (1.3) | 6(0.8) |
| COVID-19 | 13 (5.4) | 3 (0.4) |
| Musculoskeletal and connective tissue disorders | 18 (7.5) | 42 (5.8) |
| Back pain | 5 (2.1) | 16 (2.2) |
| Myalgia | 4 (1.7) | 6 (0.8) |
| Arthralgia | 3 (1.3) | 5 (0.7) |
| Neck pain | 4 (1.7) | 3 (0.4) |
| Skin and subcutaneous tissue disorders | 8 (3.3) | 32 (4.4) |
| Pruritus | 2 (0.8) | 9 (1.2) |
| Injury,­ poisoning, and procedural complications | 10 (4.2) | 30 (4.1) |
| Contusion | 3 (1.3) | 3 (0.4) |
| Investigations | 12 (5.0) | 23 (3.2) |
| Blood pressure increased | 5 (2.1) | 9 (1.2) |
| Psychiatric disorders | 4 (1.7) | 11 (1.5) |
| Nervousness | 3 (1.3) | 0 |
| Renal and urinary disorders | 0 | 10 (1.4) |
| Vascular disorders | 9 (3.8) | 10 (1.4) |
| Hypertension | 7 (2.9) | 4 (0.5) |
| Reproductive system and breast disorders | 3 (1.3) | 6 (0.8) |
| Blood and lymphatic system disorders | 3 (1.3) | 4 (0.5) |
| Metabolism and nutrition disorders | 5 (2.1) | 4 (0.5) |

Abbreviations: COVID-19, coronavirus disease 2019; Q4W, every 4 weeks; SC, subcutaneous; TEAE, treatment-emergent adverse event.

Medical Dictionary of Regulatory Activities (Version 23.1) coding dictionary applied. A subject who reported 2 or more TEAEs with the same preferred term is counted only once for that term. Included are events that were reported before the COVID-19 vaccination date, if any.

Supplementary Table 9. Subjects with Injection-Site Reaction Symptoms by Duration and Preferred Term during the Treatment Period

|  | Placebo (n = 240) | REGEN-COV  1200 mg SC Q4W (n = 729) |
| --- | --- | --- |
| Subjects with at least 1 injection site reaction symptom, n (%)^a^ | 40 (16.7) | 266 (36.5) |
| Overall time to resolution (days)^b^ |  |  |
| n | 40 | 266 |
| Mean (SD) | 5.9 (7.4) | 4.4 (6.7) |
| Median | 2.5 | 1.9 |
| Q1 : Q3 | 0.2:11.0 | 0.8:5.2 |
| Min : Max | 0.05:27.9 | 0.001:42.0 |
| Number of subjects with injection site reaction resolved in, n (%) |  |  |
| <1 day | 14 (35.0) | 97 (36.5) |
| ≥1 to <2 days | 5 (12.5) | 41 (15.4) |
| ≥2 to <3 days | 2 (5.0) | 29 (10.9) |
| ≥3 to <4 days | 4 (10.0) | 11 (4.1) |
| ≥4 to <5 days | 1 (2.5) | 18 (6.8) |
| ≥5 days | 14 (35.0) | 70 (26.3) |
| Preferred term, n (%) | | |
| Erythema | 17 (7.1) | 201 (27.6) |
| Pruritus | 2 (0.8) | 95 (13.0) |
| Nodule | 3 (1.3) | 89 (12.2) |
| Edema | 2 (0.8) | 73 (10.0) |
| Ecchymosis | 14 (5.8) | 49 (6.7) |
| Pain | 5 (2.1) | 35 (4.8) |
| Tenderness | 3 (1.3) | 24 (3.3) |
| Fibrosis | 0 | 7 (1.0) |
| Urticaria | 1 (0.4) | 4 (0.5) |
| Rash | 0 | 2 (0.3) |
| Abdominal tenderness | 0 | 1 (0.1) |
| Inflammation | 1 (0.4) | 1 (0.1) |
| Injection-site warmth | 0 | 1 (0.1) |
| Papule | 0 | 1 (0.1) |
| Paresthesia | 0 | 1 (0.1) |
| Rash macular | 0 | 1 (0.1) |
| Skin abrasion | 0 | 1 (0.1) |
| Skin hypopigmentation | 0 | 1 (0.1) |

Abbreviations: COVID-19, coronavirus disease 2019; Q4W, every 4 weeks; SC, subcutaneous; SD, standard deviation.

^a^Included are events that were reported before the COVID-19 vaccination date, if any. Only events that occurred ≥1% in the REGEN-COV group are included.

^b^For each dosing visit, if a subject had multiple injection-site reactions, time to resolution duration (days) is defined as the time from the first injection-site reaction to time that the last injection-site reaction ended.

Supplementary Table 10. Injection-Site Reactions by Study Site

|  | Placebo | REGEN-COV 1200 mg SC Q4W |
| --- | --- | --- |
| All sites combined, N | 240 | 729 |
| Subjects with at least 1 ISR, n (%) | 40 (16.7) | 266 (36.5) |
| Mean number of visits subject has experienced ISRs | 1.2 | 2.3 |
| Site 1, N | 47 | 135 |
| Subjects with at least 1 ISR, n (%) | 11 (23.4) | 88 (65.2) |
| Mean number of visits subject have experienced ISRs | 1.0 | 2.6 |
| Site 2, N | 50 | 145 |
| Subjects with at least 1 ISR, n (%) | 20 (40.0) | 104 (71.7) |
| Mean number of visits subject have experienced ISRs | 1.3 | 2.5 |
| Site 3, N | 48 | 94 |
| Subjects with at least 1 ISR, n (%) | 4 (8.3) | 12 (12.8) |
| Mean number of visits subject have experienced ISRs | 1.5 | 1.4 |
| Site 4, N | 32 | 116 |
| Subjects with at least 1 ISR | 0 | 9 (7.8) |
| Mean number of visits subject have experienced ISRs |  | 1.2 |
| Site 5, N | 29 | 101 |
| Subjects with at least 1 ISR, n (%) | 4 (13.8) | 23 (22.8) |
| Mean number of visits subject have experienced ISRs | 1.0 | 1.6 |
| Site 6, N | 16 | 64 |
| Subjects with at least 1 ISR, n (%) | 1 (6.3) | 27 (42.2) |
| Mean number of visits subject have experienced ISRs | 2.00 | 2.2 |
| Site 7, N | 18 | 74 |
| Subjects with at least 1 ISR, n (%) | 0 | 3 (4.1) |
| Mean number of visits subject have experienced ISRs |  | 1.0 |

Abbreviations: ISR, injection-site reaction; Q4W, every 4 weeks; SC, subcutaneous; TEAE, treatment-emergent adverse event.

Medical Dictionary of Regulatory Activities (Version 24.0) coding dictionary applied.

Supplementary Table 11. Summary of COVID-19 Vaccinations

|  | Placebo n = 240 | REGEN-COV 1200 mg SC Q4W  n = 729 | Total N = 969 |
| --- | --- | --- | --- |
| Subjects with COVID-19 vaccinations during the study, n (%) | | | |
| Any COVID-19 vaccine | 98 (40.8) | 256 (35.1) | 354 (36.5) |
| Moderna | 42 (17.5) | 97 (13.3) | 139 (14.3) |
| Pfizer | 52 (21.7) | 137 (18.8) | 189 (19.5) |
| Janssen | 4 (1.7) | 22 (3.0) | 26 (2.7) |
| Days from the last administration of study drug to receive the first dose of COVID-19 vaccination^a^ | | | |
| n | 98 | 256 | 354 |
| Mean (SD) | 58.4 (43.6) | 69.1 (41.4) | 66.1 (42.2) |
| Median | 57.0 | 90.0 | 77.0 |
| Q1 : Q3 | 18.0 : 97.0 | 28.0 : 100.0 | 25.0 : 99.0 |
| Min : Max | –20.0 : 194.0 | -11.0 : 186.0 | –20.0 : 194.0 |
| Days of follow-up post the first dose of COVID-19 vaccination^b^ | | | |
| n | 98 | 256 | 354 |
| Mean (SD) | 66.5 (35.54) | 56.3 (35.14) | 59.1 (35.50) |
| Median | 56.5 | 47.0 | 51.0 |
| Q1 : Q3 | 42.0 : 96.0 | 33.5 : 74.0 | 36.0 : 78.0 |
| Min : Max | 3.0 : 156.0 | 1.0 : 155.0 | 1.0 : 156.0 |

Abbreviations: COVID-19, coronavirus disease 2019; Q4W, every 4 weeks; SC, subcutaneous; SD, standard deviation.

^a^The days from the last administration of study drug are calculated as the date of first dose of COVID-19 vaccination minus the date of last study drug administration plus 1. If the date of last study drug administration is before the date of first dose of COVID-19 vaccination, then the days from the last administration of study drug are calculated as the date of first dose of COVID-19 vaccination minus the date of last study drug administration.

**^b^**The days of follow-up post the first dose of COVID-19 vaccination are calculated as the earliest date of end of study, death, or the data cut-off date minus the date of first dose of COVID-19 vaccination plus 1. If the end of study date is before the date of first dose of COVID-19 vaccination, then the days of follow-up are calculated as the date of end of study minus the date of first dose of COVID-19 vaccination.

Supplementary Table 12. Overview of Adverse Events After Subjects Received COVID-19 Vaccination

|  | Placebo (n = 98) | | REGEN-COV 1200 mg SC Q4W  (n = 256) |
| --- | --- | --- | --- |
| Number of TEAEs | 36 | | 72 |
| Number of grade ≥3TEAEs | 0 | | 1 |
| Number of serious TEAEs | 0 | | 1 |
| Number of AESIs | 0 | | 0 |
| Number of TEAEs resulting in study drug being withdrawn | 1 | | 0 |
| Number of TEAEs resulting in death | 0 | | 0 |
| Subjects with at least 1 TEAE, n (%) | 18 (18.4) | | 39 (15.2) |
| Subjects with at least 1 grade ≥3 TEAE^a^, n (%) | 0 | | 1 (0.4) |
| Subjects with at least 1 serious TEAE, n (%) | 0 | | 1 (0.4) |
| Subjects with at least 1 AESI, n (%) | 0 | | 0 |
| Subjects with at least 1 TEAE resulting in study drug being withdrawn, n (%) | 1 (1.0) | | 0 |
| Subjects with any TEAE resulting in death, n (%) | 0 | | 0 |
| Preferred term, n (%) | | | |
| General disorders and administration site conditions, n (%) | 8 (8.2) | 22 (8.6) | |
| Vaccination-site pain | 7 (7.1) | 20 (7.8) | |
| Chills | 0 | 1 (0.4) | |
| Fatigue | 1 (1.0) | 1 (0.4) | |
| Non-cardiac chest pain | 0 | 1 (0.4) | |
| Pyrexia | 0 | 1 (0.4) | |
| Vaccination-site bruising | 0 | 1 (0.4) | |
| Pain | 1 (1.0) | 0 | |
| Injury,­ poisoning, and procedural complications | 10 (10.2) | 15 (5.9) | |
| Vaccination complication | 9 (9.2) | 15 (5.9) | |
| Procedural pain | 1 (1.0) | 0 | |
| Musculoskeletal and connective tissue disorders | 1 (1.0) | 4 (1.6) | |
| Musculoskeletal stiffness | 0 | 1 (0.4) | |
| Neck pain | 0 | 1 (0.4) | |
| Osteoporosis | 0 | 1 (0.4) | |
| Pain in extremity | 1 (1.0) | 1 (0.4) | |
| Nervous system disorders | 1 (1.0) | 3 (1.2) | |
| Dizziness | 0 | 1 (0.4) | |
| Headache | 1 (1.0) | 1 (0.4) | |

Abbreviations: AESI, adverse event of special interest; COVID-19, coronavirus disease 2019; Q4W, every 4 weeks; SC, subcutaneous; TEAE, treatment-emergent adverse event.

Medical Dictionary of Regulatory Activities (Version 24.0) coding dictionary applied.

^a^A single grade 3 TEAE occurred following COVID-19 vaccination: brain mass frontal lobe. This TEAE was assessed as not related to study drug.

Supplementary Table 13. Pharmacokinetics of the Concentrations of Casirivimab and Imdevimab in Serum over Time in Subjects Receiving REGEN-COV with SARS-CoV-2 Infection

| Nominal Sampling (Days) | Concentrations in Serum (mg/L) | | | | | |
| --- | --- | --- | --- | --- | --- | --- |
|  | Casirivimab 600 mg SC (N = 3) | | Imdevimab 600 mg SC (N = 3) | | Casirivimab+imdevimab (REGEN-COV) 1200 mg SC Q4W (N = 3) | |
|  | n | Mean (SD) | n | Mean (SD) | n | Mean (SD) |
| 0 | 3 | 0 (0) | 3 | 0 (0) | 3 | 0 (0) |
| 7 | 3 | 57.0 (10.6) | 3 | 54.3 (8.81) | 3 | 111 (17.6) |
| 28 | 3 | 38.0 (11.7) | 3 | 30.3 (11.1) | 3 | 68.3 (22.7) |
| 56 | 3 | 57.7 (23.3) | 3 | 43.7 (19.9) | 3 | 101 (43.1) |
| 84 | 1 | 56.5 (---) | 1 | 39.9 (---) | 1 | 96.4 (---) |
| 112 | 2 | 51.8 (---) | 2 | 35.0 (---) | 2 | 86.7 (---) |
| 140 | 1 | 54.1 (---) | 1 | 35.6 (---) | 1 | 89.7 (---) |
| 168 | 2 | 79.5 (---) | 2 | 56.4 (---) | 2 | 136 (---) |
| 224 | 2 | 25.7 (---) | 2 | 18.2 (---) | 2 | 43.9 (---) |

Abbreviations: COVID-19, coronavirus disease 2019; Q4W, every 4 weeks; SC, subcutaneous; SD, standard deviation.

N is equal to total number of subjects; n is equal to number of subjects contributing to each timepoint. Combined concentrations (REGEN-COV) were calculated only when both the analytes were not missing. There are no virology data to confirm these 3 COVID-19 diagnoses, which were recorded as adverse events by the investigator on the basis of clinical assessment.

Supplementary Figure 1. Study Design

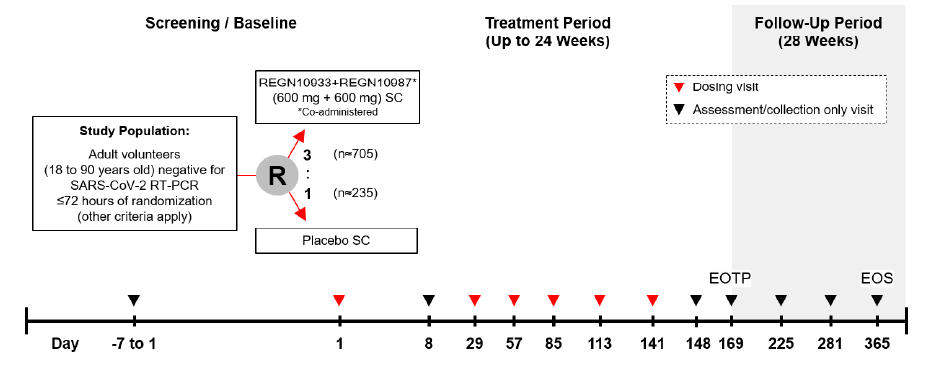

Abbreviations: EOS; end of study; EOTP, end-of-treatment period; R, randomized; RT-PCR, reverse transcription polymerase chain reaction; SARS-CoV-2, severe acute respiratory syndrome coronavirus 2; SC, subcutaneous.

The study design is depicted from day –7 to day 365 (EOS). Study dug was given as up to 6monthly administrations on study days 1, 29, 57, 85, 113, and 141. The EOTP visit occurred 1 month after the last administration of the study drug, after which subjects entered the 28-week follow-up period leading to EOS on day 365. As of the data cut-off of May 21, 2021, all eligible subjects had completed the end-of-treatment visit and follow-up was ongoing, with not all subjects having completed the entire study.

Supplementary Figure 2. SARS-CoV-2 Infection by Calendar Day During the Treatment Period

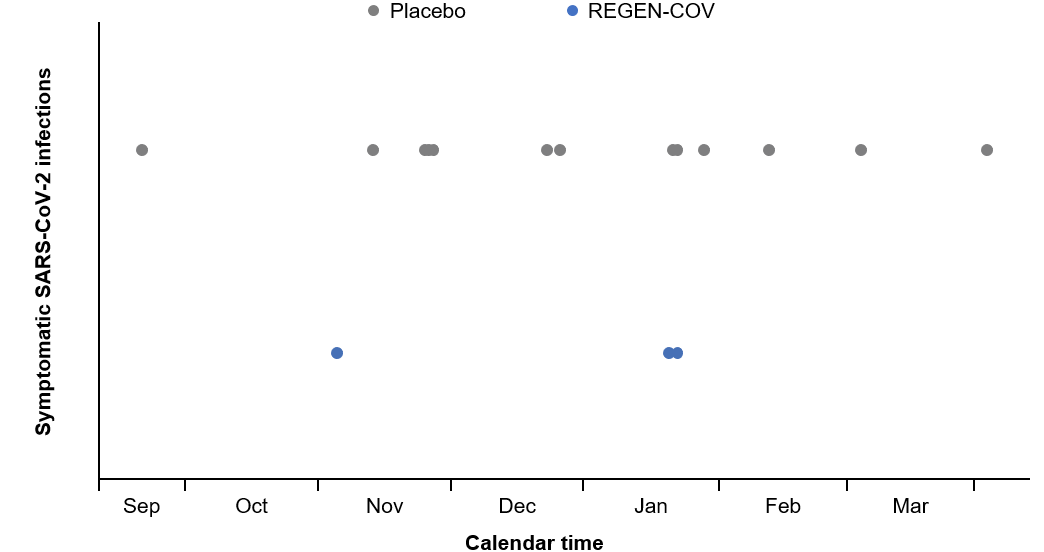

Abbreviations: COVID-19, coronavirus disease 2019; SARS-CoV-2, severe acute respiratory syndrome coronavirus 2.

Individual symptomatic SARS-CoV-2 (COVID-19) cases are depicted as dots by calendar time in months from year 2020 to 2021. Symptomatic SARS-CoV-2 determination was made by the investigator on the basis of clinical assessment.

Supplementary Figure 3. Injection-Site Reaction Frequency by (A) Dose (All 7 Sites Combined), (B) Study Site, and (C) Dose (Sites 1 and 2 Removed)

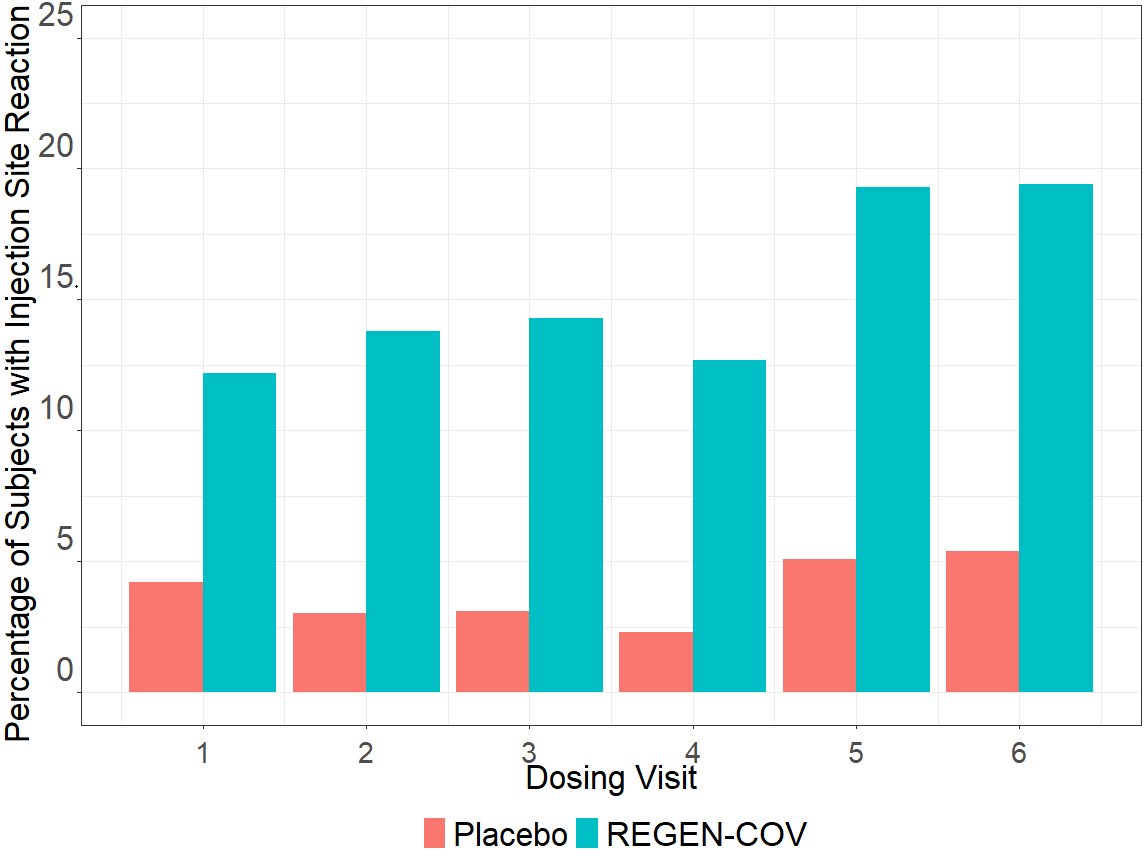

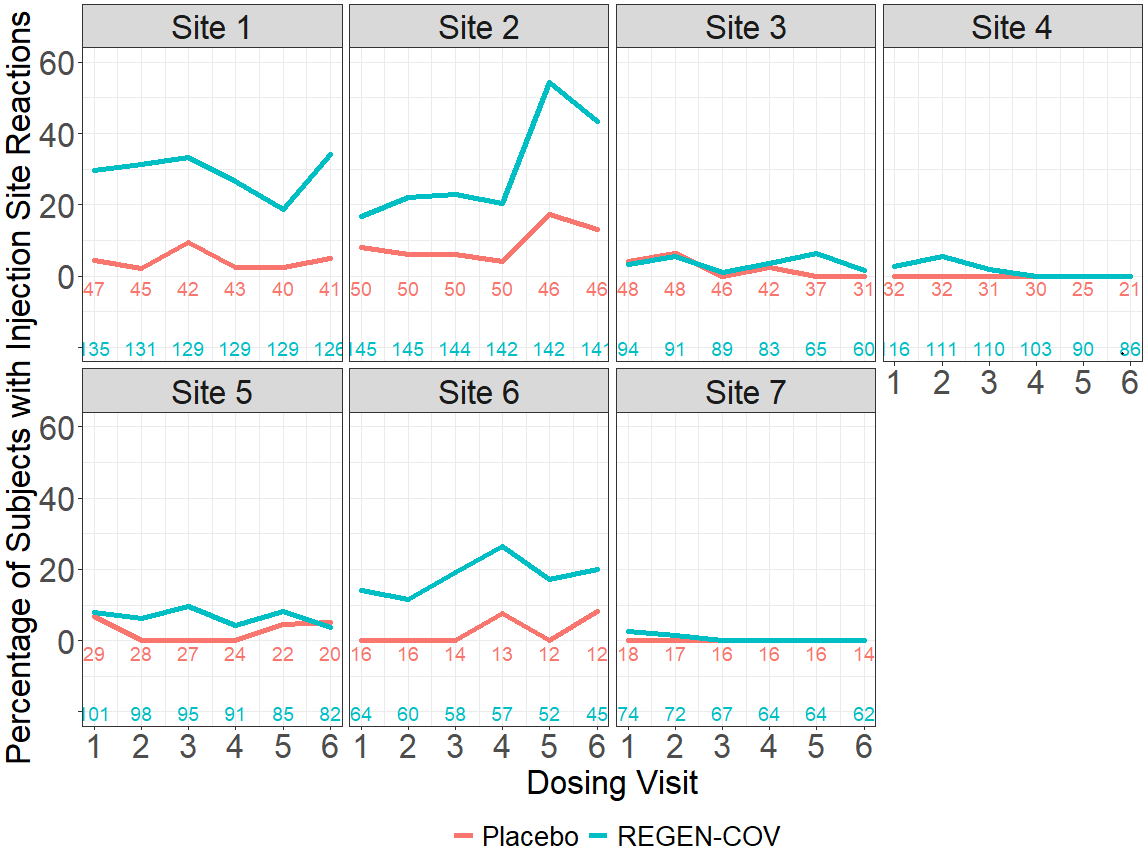

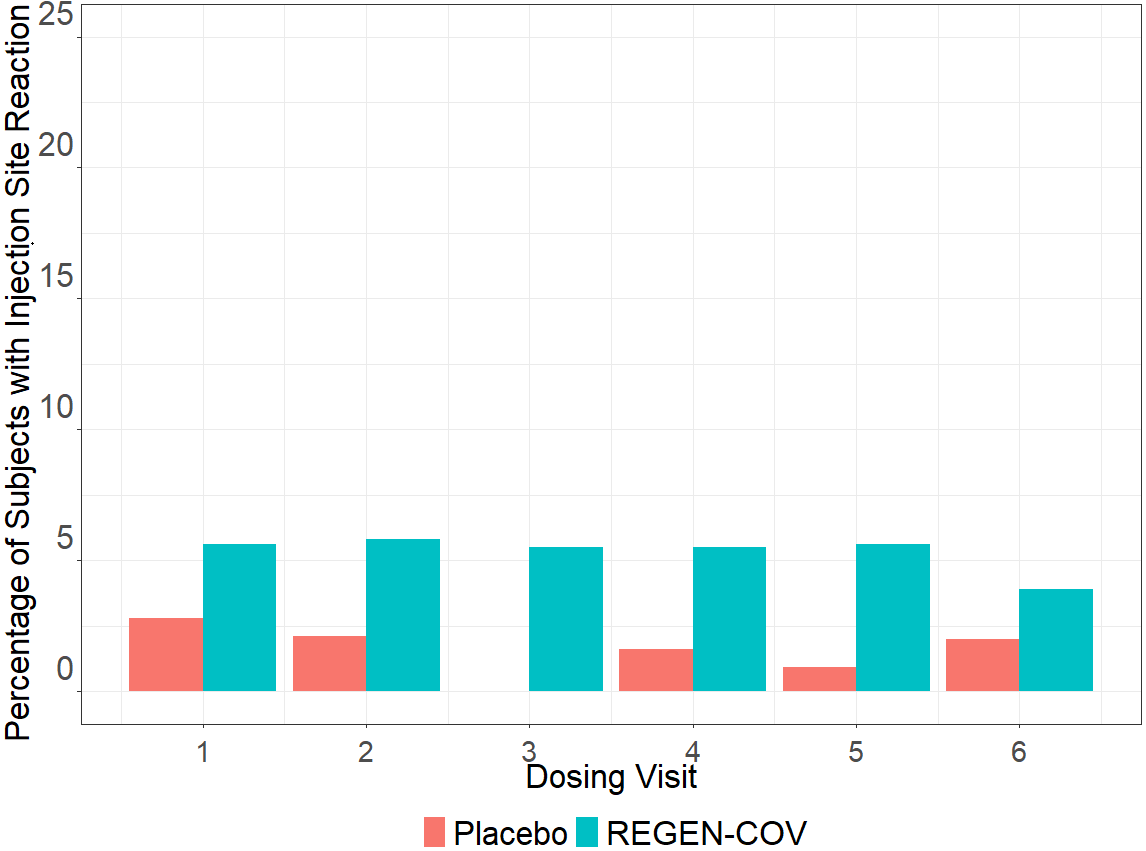

Panel A displays injection-site reactions by site over the course of 6 dose administrations of REGEN-COV or placebo. Panel B displays the averaged combined incidence of injection-site reactions across all 7 sites. Panel C displays the same data as panel B but with the outlier sites (sites 1 and 2) with the highest rate of injection-site reactions removed.

Supplementary Figure 4. Concentrations of (A) Total Casirivimab and (B) Total Imdevimab in Serum Over Time by Negative or Positive Anti-Drug Antibody Status

**A.**

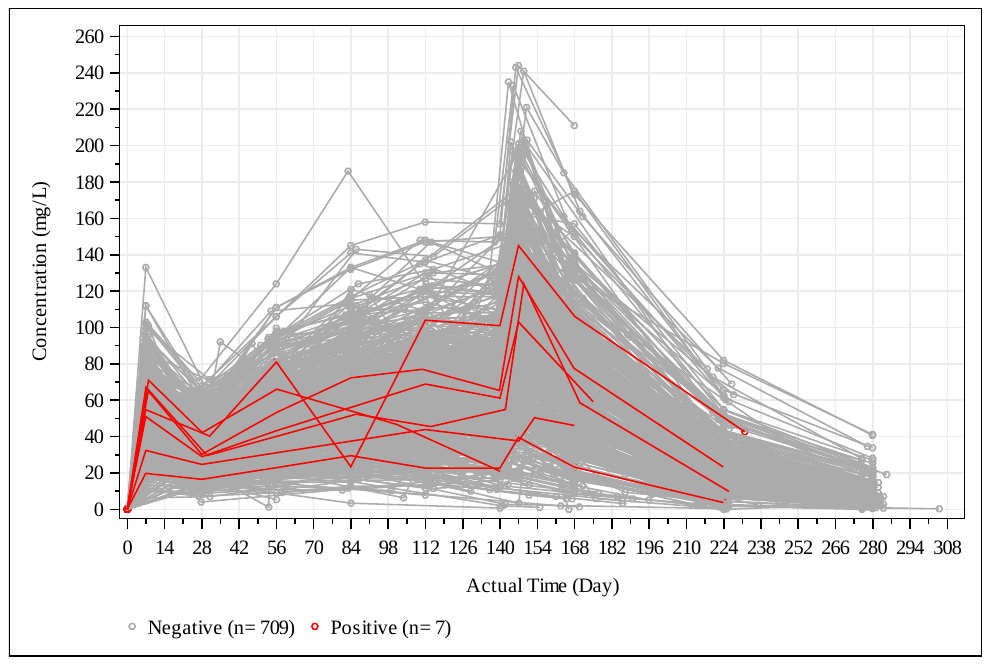

**B.**

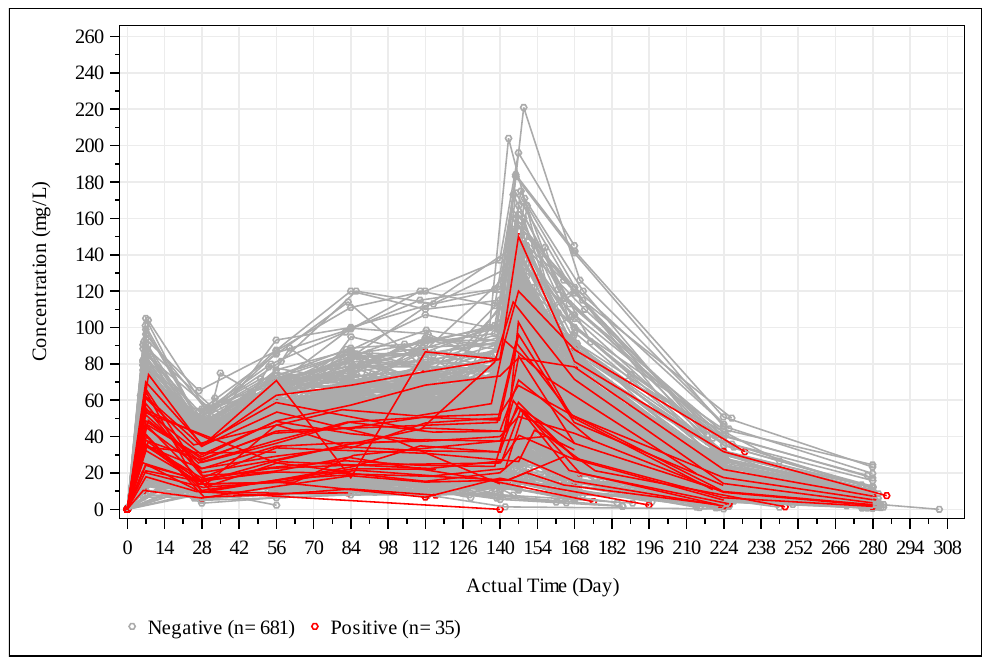

n is equal to the number of subjects contributing to each category.
